# Diagnostic accuracy of cerebrospinal fluid and blood biomarkers for the differential diagnosis of sporadic Creutzfeldt-Jakob disease: a (network) meta-analysis

**DOI:** 10.1101/2021.03.25.21254312

**Authors:** Nicole Rübsamen, Stephanie Pape, Stefan Konigorski, Antonia Zapf, Gerta Rücker, André Karch

## Abstract

**Objective:** To conduct a systematic review of cerebrospinal fluid (CSF) and blood biomarkers as diagnostic tests for sporadic Creutzfeldt-Jakob disease (sCJD) in a specialised care setting and to compare diagnostic accuracies in a network meta-analysis (NMA).

**Methods:** We searched Medline, Embase, and the Cochrane Library for diagnostic studies of sCJD biomarkers. Risk of bias was assessed with the QUADAS-2 tool. We used a generalised bivariate model to conduct individual biomarker meta-analyses, and to estimate between-study variability. To investigate sources of heterogeneity, we performed subgroup analyses based on QUADAS-2 quality and clinical criteria. For the NMA, we applied a Bayesian beta-binomial ANOVA model. The study protocol was registered at PROSPERO (CRD42019118830).

**Results:** Out of 2,976 publications screened, we included 16 studies, which investigated 14-3-3β Western blot (n=13), 14-3-3γ ELISA (n=3), NfL (n=1), NSE (n=1), p-tau181/t-tau ratio (n=2), RT-QuIC (n=6), S100B (n=3), t-tau (n=12), and t-tau/Aβ42 ratio (n=1) in CSF. No included study investigated blood biomarkers. Many diagnostic studies excluded had strong limitations in study design. In the NMA, RT-QuIC (0.93; 95% CI [0.87, 0.96]) and NfL (0.94 [0.81, 0.99]) were the most sensitive biomarkers. RT-QuIC was the most specific biomarker (0.96 [0.86, 0.99]), and had the highest balanced accuracy (0.94). Heterogeneity in accuracy estimates was high between studies, especially for specificity.

**Conclusions:** Our NMA identified RT-QuIC as the overall most accurate biomarker, partially confirming current guidelines. The severe shortcomings identified in many diagnostic studies for sCJD biomarkers need to be addressed in future studies in the field.

## TEXT

### Introduction

The sporadic form of Creutzfeldt-Jakob disease (sCJD) is the world’s most common human prion disease with an incidence of about 1–2 cases per million and year.^1,2^ The reference standard for the definite diagnosis of sCJD is the post-mortem neuropathological examination of the brain, which is of little benefit to patients during their lifetime. An alternative is the use of a diagnostic composite reference standard.^3,4^ Based on clinical criteria combined with either defined changes in magnetic resonance imaging (MRI), a characteristic electroencephalogram (EEG), or a positive biomarker test, patients are differentiated into non-sCJD patients as well as probable and possible sCJD cases.

Detection of the proteins 14-3-3 and the PrPSc aggregation assay RT-QuIC (Real-Time Quaking Induced Conversion) are currently incorporated in the diagnostic composite reference standard.^3,5^ Other biomarkers, e.g. tau (total tau [t-tau] or phosphorylated tau [p-tau]), neuron-specific enolase (NSE), neurofilament light chain (NfL), and S100B, have been proposed as an addition or replacement option for the biomarkers in the composite criteria for the differential diagnosis of sCJD. There is no evidence from systematic reviews and diagnostic meta-analyses that have performed a comparative investigation of the accuracy of biomarkers suitable for the differential diagnosis of sCJD. Previous reviews in the field focused either on a single biomarker^6^, or did not formally compare biomarkers in a meta-analysis.^7^ We, therefore, conducted a network meta-analysis based on a systematic review to compare the accuracy of established biomarker tests measured in the blood or cerebrospinal fluid (CSF) to diagnose sCJD in a specialised care setting under real-life conditions.

## Methods

### Search strategy and selection criteria

For this systematic review and meta-analysis, we systematically searched Medline, Embase, and the Cochrane Library for diagnostic studies that assessed the accuracy of blood or CSF biomarkers to diagnose sCJD (Table e-1). One author (SP) searched all databases on 25^th^ July 2018 (initial search) and 23^rd^ September 2020 (update). The search term for Medline was (((((Biomarker) OR biomarkers [MeSH Terms])) OR ((Diagnosis) OR diagnosis [MeSH Terms]))) AND ((((Creutzfeldt Jakob disease) OR CJD)) OR cjd creutzfeldt jakob disease [MeSH Terms]) AND ((CSF OR cerebrospinal) OR (blood or serum or plasma)). SP merged all search results and removed duplicates.

Studies were included if they assessed blood or CSF biomarkers’ accuracy for the differentiation of sCJD from other diseases in a specialised care setting (Table e-2). Eligible studies had to use established diagnostic criteria of sCJD^3,4^ and established diagnostic criteria for diseases in the non-CJD groups (e.g., Alzheimer’s disease, vascular dementia, frontotemporal dementia, dementia with Lewy bodies, alcohol-induced dementia) as reference standard. The non-CJD group had to represent patients suspected as a CJD case and referred to the specialised care centre for a first diagnostic workup. The biomarker tests had to be performed during the patient’s first diagnostic workup for her/his symptoms without a pre-defined diagnosis. Furthermore, the studies had to provide sufficient information to construct a diagnostic contingency table (i.e., true and false positives and negatives). There were no constraints regarding time, language, stage of sCJD, patient population, type of biomarker, reference standard, or study design. Different restrictions were used for specific biomarkers: Results for 14-3-3 and RT-QuIC were included if there was no risk of incorporation bias,^8^ i.e. if 14-3-3 or RT-QuIC were not incorporated in the reference standard used. Results for t-tau were included if the cut-off used to classify a test result as positive/negative was within ± 10% of the recommended cut-off 1300 pg/mL.^9^ For all other biomarkers, the cut-off values used in the study needed to be based on external knowledge or training datasets. Methods used for measurement of S100B differed between studies. We allowed, therefore, for different cut-off values as long as they fulfilled the criteria above.

However, we excluded measurements of RT-QuIC using the first-generation assay (PQ-CSF) because this technology is no longer used in clinical practice. Only measurements of RT-QuIC using the second-generation assay (IQ-CSF) were included.

Two authors (SP and AK) independently reviewed all titles, abstracts, and, if publications were included based on this information, full texts. Discrepancies were resolved in a consensus meeting. An agreement was reached regarding all discrepancies so that referral to a third reviewer was not necessary.

All reference lists of publications included in the final systematic review were searched for additional studies that were missed, and these studies were further included in the review. Moreover, we discussed with experts from sCJD-specialised care centres if the search strategy missed studies known to them. We did not specifically assess grey literature sources in addition to this strategy because due to the rarity of sCJD and the clustering of patients in few specialised care settings worldwide, newly obtained findings are generally made available in manuscript form.

The study protocol and statistical analysis plan were registered with PROSPERO on January 9^th^, 2019 (CRD42019118830) after the beginning of preliminary searches, piloting of the study selection process, and formal screening of search results against eligibility criteria; however, before data extraction, risk of bias (quality) assessment and data analysis had been started.

### Data extraction and quality assessment

SP extracted clinical and demographic information (number of patients, clinical diagnosis, age, sex, setting, recruitment, and sampling procedure) as well as details of the assays and cut-offs used. Two authors (NR and SP) independently extracted the number of true and false positives and negatives (stratified by the level of certainty of sCJD diagnosis). An agreement was reached regarding all discrepancies. We contacted the corresponding authors if further information was needed.

SP, AK, and NR performed their risk of bias assessments for the included studies independently using the Quality Assessment of Diagnostic Accuracy Studies-2 (QUADAS-2) tool, the only validated tool for estimating risk of bias in diagnostic studies.^10^ QUADAS-2 covers quality evaluations in four different domains (patient selection, index test, reference standard, flow and timing) by using signalling questions related to the research question, and is supplemented by applicability inquiries. AK and SP first piloted the tool by using two randomly selected publications.^9,11^ Satisfactory agreement was reached in all domains so that the QUADAS-2 signalling questions were retained unchanged for the assessment of the included studies (Table e-3).

### Data analysis

All analyses were performed with R version 4.0.3.^12^

#### Systematic review

We calculated Krippendorff’s alpha^13^ to assess the reliability of the risk of bias assessment between the investigators.

#### Meta-analyses of individual biomarkers

We tabulated true positives, false negatives, false positives, and true negatives in patients with and without sCJD, stratified by study and level of certainty of sCJD diagnosis. Based on this, we calculated estimates of sensitivity and specificity and their 95% confidence intervals (CI). To investigate publication bias, we constructed funnel plots of effective sample size versus the estimated log diagnostic odds ratios and did a regression test of asymmetry.^14^ To synthesise data, we implemented the generalised linear mixed model approach by Chu and Cole,^15^ which uses a bivariate binomial model to jointly analyse pairs of sensitivity and specificity, using the *glmer* function in the R package *lme4* (version 1.1-25).^16^ This approach allows estimating the correlation between true positive rate (TPR) and false-positive rate (FPR) as well as the between-study standard deviation (SD) for both of them via random effects, which provides information on the heterogeneity of the results.^17^ To investigate sources of heterogeneity, we performed subgroup analyses based on QUADAS-2 quality and clinical criteria. Although there exist approaches for meta-analyses of diagnostic test accuracy studies with multiple cut-offs, we decided against using them because biomarker cut-offs were either homogeneous or were applied to different technical platforms, so that different measurement scales were used. We repeated all analyses for different levels of certainty of sCJD diagnosis: 1) definite sCJD cases, 2) definite and probable sCJD cases, and 3) definite, probable, and possible sCJD cases.

#### Network meta-analysis

To compare the diagnostic accuracy of different biomarkers, we applied a diagnostic network meta-analysis (NMA) approach for evidence synthesis. We used the beta-binomial analysis of variance model for NMA of diagnostic test accuracy data as described by Nyaga et al.^18^ to combine direct and indirect evidence simultaneously. This arm-based generalised linear mixed model models sensitivity and specificity as repeated measures jointly through a copula function and assumes that the missing tests/arms are missing at random. The models were fitted in the Bayesian framework with beta(1,1) = U(0,1) as prior distribution on the hyper-parameters using Stan^19^ through the R package *rstan* (version 2.21.2).^20^ We repeated the network meta-analyses for different levels of certainty of sCJD diagnosis (as described above).

### Data Availability Statement

All data generated or analysed during this study are included in this article.

## Results

### Systematic review

Our database search retrieved 2,976 articles. After reviewing the titles and abstracts, we excluded 2,668 articles, mostly because these studies were not diagnostic studies, they duplicated data already included, or the case definition or study population was not appropriate. After a full-text review of the remaining 308 articles, we found that 86 studies were notdiagnostic studies and that 32 articles reported duplicated data. Among the remaining 190 diagnostic studies, we excluded 171 studies because of inappropriate study populations or missing information on key study characteristics, leaving 19 articles for risk of bias assessment (Figure 1).

**Figure 1.**
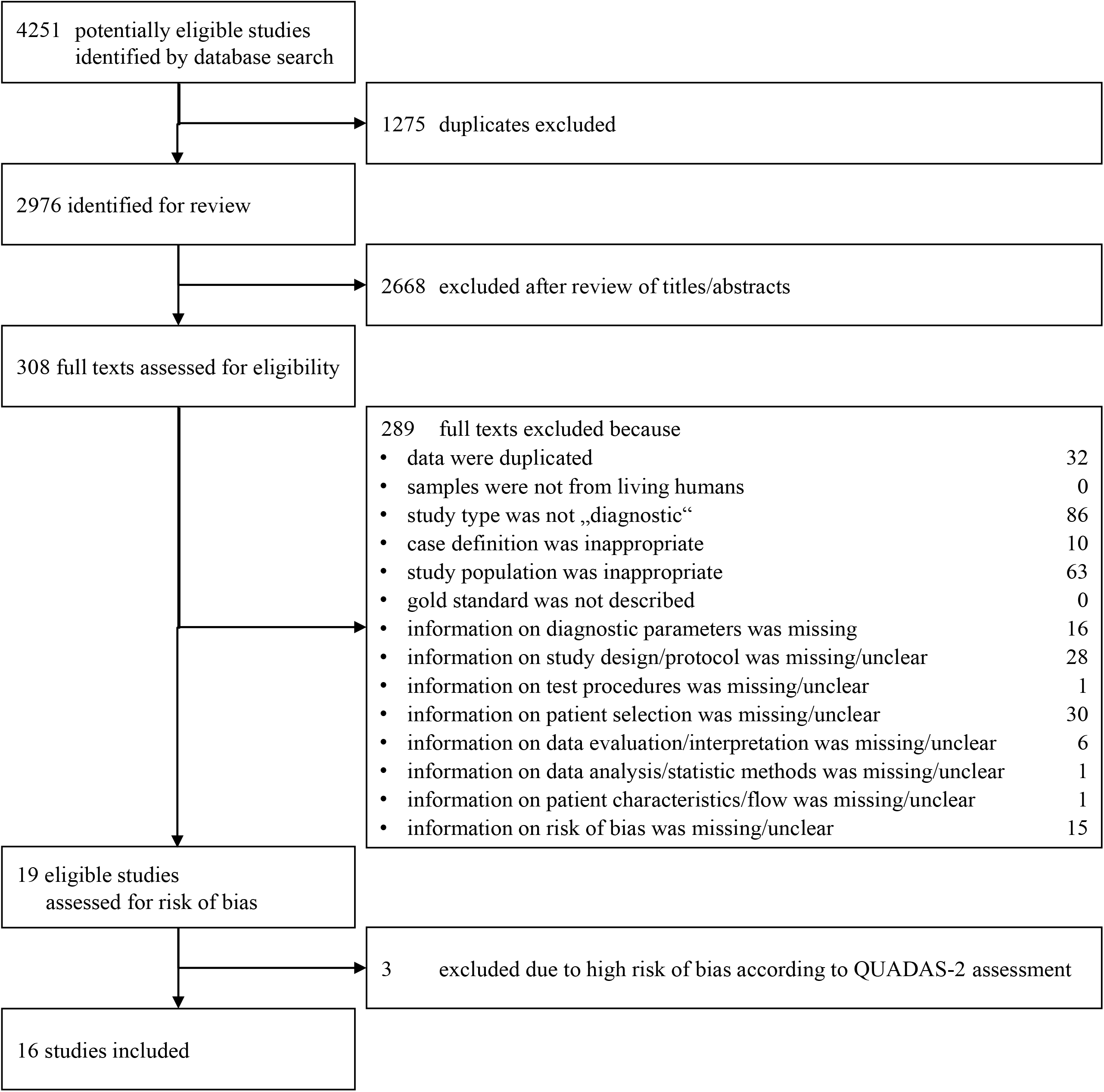
Flow chart of study selection

Sufficient inter-rater reliability was achieved in the core areas of patient selection, blinding and missing values, which are particularly important for quality assessment (Table e-4). The reviewers discussed all 19 studies together, and differences in the risk of bias assessment (which were mainly based on high vs unclear risk of bias in studies where specific features were only partially reported) could be fully resolved (Table e-4). Based on high or unclear risks of bias and poor applicability, the studies by Hamlin et al.,^11^ Rudge et al.,^21^ and Wang et al.^22^ were excluded, leaving 16 studies for further quantitative analyses^9,23–37^ (Table 1).

**Table 1:**
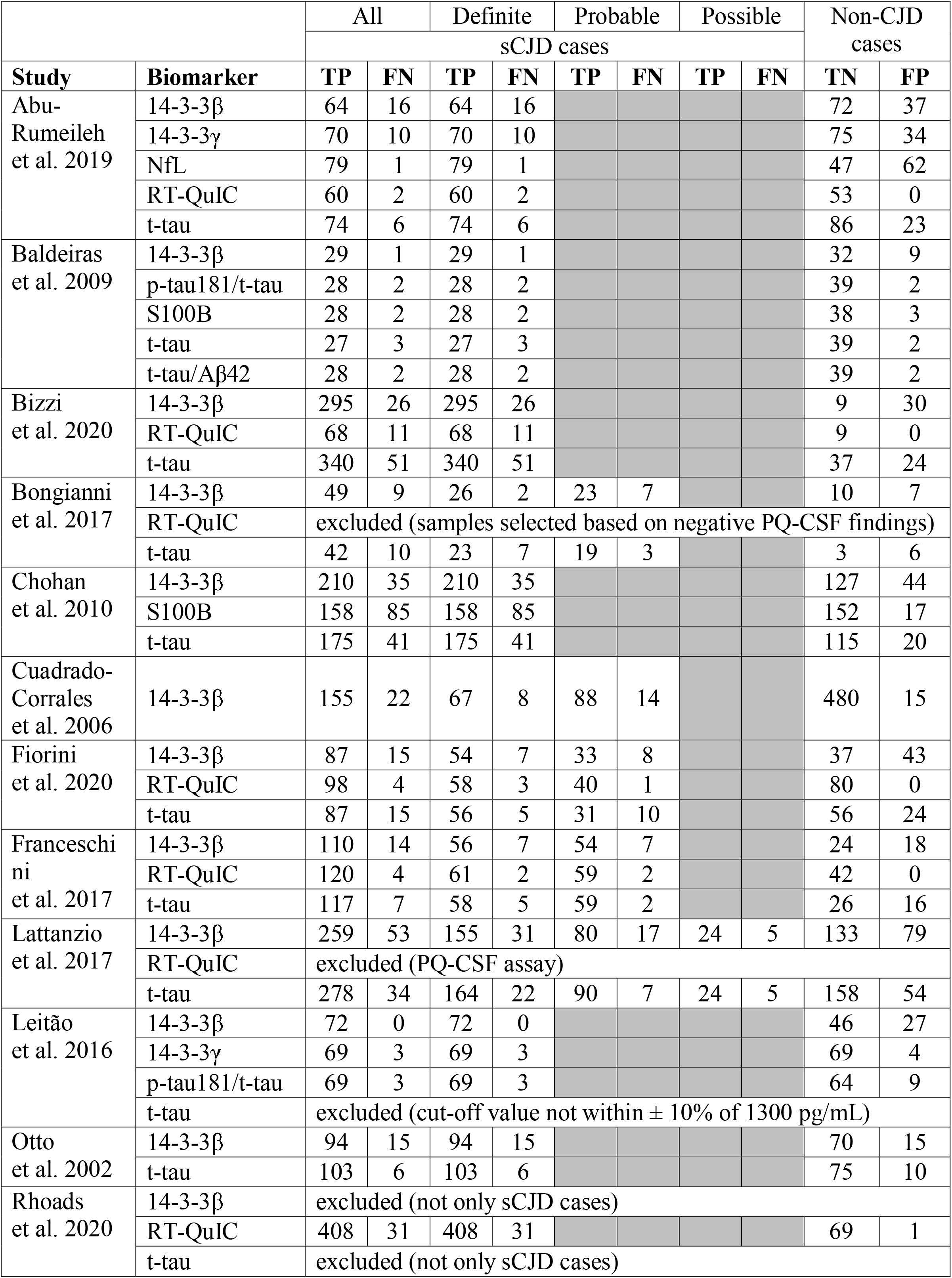
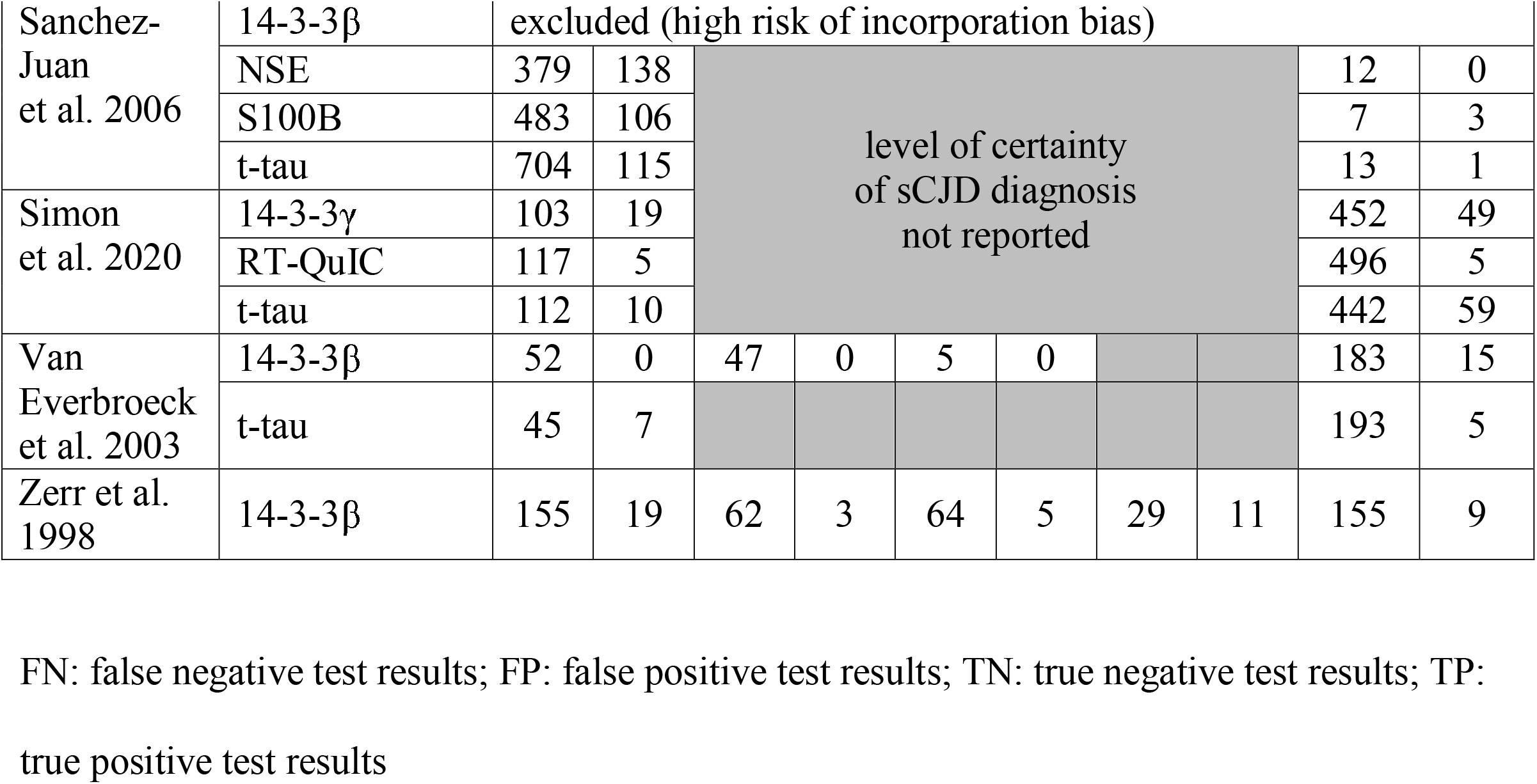
Results of diagnostic sCJD blood or CSF biomarker studies that met the inclusion criteria

Fourteen of these studies investigated 14-3-3β via Western blot. The results regarding 14-3-3β in the study by Sanchez-Juan et al.^34^ were, however, excluded due to high risk of incorporation bias, leaving 13 studies that investigated 14-3-3β. Three studies investigated 14-3-3γ via ELISA. NfL, NSE, p-tau181/t-tau ratio, RT-QuIC, S100B, and t-tau/Aβ42 ratio where investigated in one, one, two, six, three, and one studies, respectively (Table 1). Thirteen studies investigated t-tau, but the study by Leitão et al.^32^ used the cut-off 1035 pg/mL to classify a test result as positive, which was outside our pre-defined interval around the recommended cut-off of 1300 pg/mL.^9^ The results regarding tau in the study by Leitão et al.^32^ were, thus, excluded. Studies included in these analyses investigated between one and five biomarkers in the same study population, providing direct comparisons for some of the biomarkers involved.

### Meta-analyses of individual biomarkers

The regression tests of asymmetry in the funnel plots did not indicate publication bias for any studied biomarkers (Figure e-1). Among definite sCJD cases, the range of observed sensitivities was 0.80 to 0.99 for 14-3-3β, 0.88 to 0.96 for 14-3-3γ, 0.86 to 0.96 for RT-QuIC, 0.65 to 0.93 for S100B, and 0.77 to 0.94 for t-tau (Figure 2). The range of observed specificities was 0.24 to 0.97 for 14-3-3β, 0.69 to 0.95 for 14-3-3γ, 0.95 to 0.99 for RT-QuIC, 0.90 to 0.93 for S100B, and 0.33 to 0.95 for t-tau (Figure 2). Individual meta-analyses of NfL, NSE, p-tau181/t-tau ratio, and t-tau/Aβ42 ratio were not conducted due to the low number of studies. Heterogeneity, estimated via SD (TPR) and SD (FPR), was high for all individual meta-analyses so that no pooled estimates were reported (Table e-5–Table e-9, Figure e-4–Figure e-8). Ranges of observed sensitivities and specificities were similar when including lower levels of certainty of sCJD diagnosis (Figure e-2, Figure e-3) except for the specificity of S100B, which was considerably lower when all levels of certainty were included. When including definite sCJD cases only, heterogeneity among studies was highest for the sensitivity of S100B (SD (TPR): 0.91); heterogeneity in specificity was high for all biomarkers but S100B (Figure 2).

**Figure 2.**
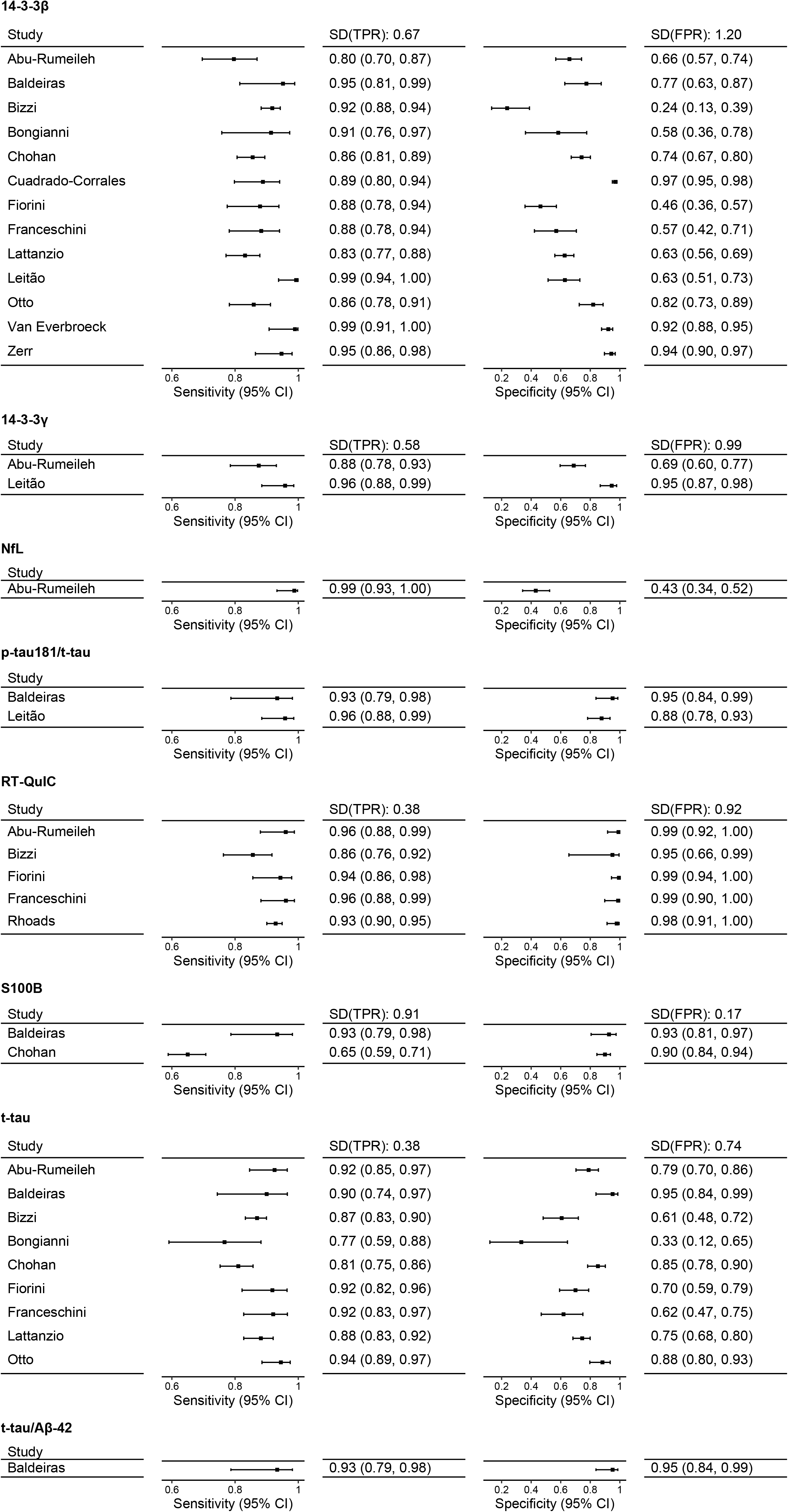
Sensitivity and specificity of CSF biomarkers for the diagnosis of definite sCJD *CI: Confidence interval; FPR: False positive rate; SD: Standard deviation; TPR: True positive rate*

There were various sources of heterogeneity for all biomarkers in all scenarios; the most important ones (identified by a decrease of heterogeneity in the respective subgroup analysis) were the definition of the study population (especially concerning the group without CJD), study design, blinding of the reference standard and clinical characteristics of the patients (Table e-5– Table e-9, Figure e-4–Figure e-8).

### Network meta-analysis

Since differences in estimates between studies in the individual meta-analyses were consistent across different biomarkers, an evidence-synthesis approach based on the combination of intra-study differences was used to derive pooled estimates of diagnostic accuracy. Figure 3 summarises the direct evidence from all studies. In the NMA analysis based on definite sCJD cases only, RT-QuIC was the most specific (0.96 [0.85, 1.00]) and the second most sensitive (0.91 [0.83, 0.96]) biomarker (Figure 4, Table 2). The balanced accuracy ([sensitivity + specificity]/2) was also highest for RT-QuIC (0.93). While NfL had the highest sensitivity in this NMA analysis (0.92 (0.72, 0.99)), its specificity was the lowest among all biomarkers (0.45 (0.15, 0.79)). Ratios involving t-tau (p-tau181/t-tau, t-tau/Aβ42) had higher accuracies than t-tau alone (Table 2); however, sensitivities and specificities were estimated with wider 95% CI (Figure 4) because fewer studies investigated these ratios than t-tau alone.

**Table 2:** Results of network meta-analyses (sorted by accuracy)

|  | Definite, probable,<br>or possible sCJD |  |  | Definite or<br>probable sCJD |  |  | Definite sCJD |  |  |
| --- | --- | --- | --- | --- | --- | --- | --- | --- | --- |
| Biomarker | Sensi-<br>tivity | Speci-<br>ficity | Accu-<br>racy | Sensi-<br>tivity | Speci-<br>ficity | Accu-<br>racy | Sensi-<br>tivity | Speci-<br>ficity | Accu-<br>racy |
| RT-QuIC | 0.93<br>(0.87,<br>0.96) | 0.96<br>(0.87,<br>0.99) | 0.94 | 0.93<br>(0.86,<br>0.96) | 0.96<br>(0.87,<br>0.99) | 0.94 | 0.91<br>(0.83,<br>0.96) | 0.96<br>(0.85,<br>1.00) | 0.93 |
| p-tau181/<br>t-tau | 0.91<br>(0.79,<br>0.98) | 0.84<br>(0.58,<br>0.96) | 0.88 | 0.91<br>(0.76,<br>0.98) | 0.84<br>(0.59,<br>0.96) | 0.88 | 0.90<br>(0.73,<br>0.98) | 0.83<br>(0.58,<br>0.96) | 0.88 |
| t-tau/<br>Aβ42 | 0.86<br>(0.64,<br>0.97) | 0.82<br>(0.47,<br>0.98) | 0.84 | 0.87<br>(0.63,<br>0.98) | 0.83<br>(0.48,<br>0.97) | 0.85 | 0.86<br>(0.60,<br>0.98) | 0.82<br>(0.46,<br>0.97) | 0.84 |
| 14-3-3γ | 0.87<br>(0.76,<br>0.95) | 0.80<br>(0.58,<br>0.92) | 0.83 | 0.86<br>(0.74,<br>0.94) | 0.79<br>(0.59,<br>0.93) | 0.83 | 0.88<br>(0.72,<br>0.96) | 0.75<br>(0.47,<br>0.92) | 0.82 |
| t-tau | 0.88<br>(0.84,<br>0.91) | 0.77<br>(0.66,<br>0.86) | 0.82 | 0.88<br>(0.84,<br>0.91) | 0.76<br>(0.63,<br>0.85) | 0.82 | 0.87<br>(0.81,<br>0.92) | 0.72<br>(0.58,<br>0.82) | 0.80 |
| 14-3-3β | 0.88<br>(0.84,<br>0.92) | 0.69<br>(0.58,<br>0.79) | 0.79 | 0.89<br>(0.83,<br>0.92) | 0.69<br>(0.57,<br>0.79) | 0.79 | 0.89<br>(0.84,<br>0.93) | 0.69<br>(0.57,<br>0.79) | 0.79 |
| S100B | 0.77<br>(0.64,<br>0.89) | 0.81<br>(0.58,<br>0.94) | 0.79 | 0.75<br>(0.55,<br>0.90) | 0.85<br>(0.63,<br>0.95) | 0.80 | 0.74<br>(0.53,<br>0.91) | 0.84<br>(0.59,<br>0.95) | 0.79 |
| NSE | 0.71<br>(0.50,<br>0.88) | 0.85<br>(0.52,<br>0.99) | 0.78 |  |  |  |  |  |  |
| NfL | 0.94<br>(0.80,<br>0.99) | 0.46<br>(0.17,<br>0.80) | 0.70 | 0.93<br>(0.74,<br>0.99) | 0.46<br>(0.15,<br>0.80) | 0.70 | 0.92<br>(0.72,<br>0.99) | 0.45<br>(0.15,<br>0.79) | 0.69 |

**Figure 3.**
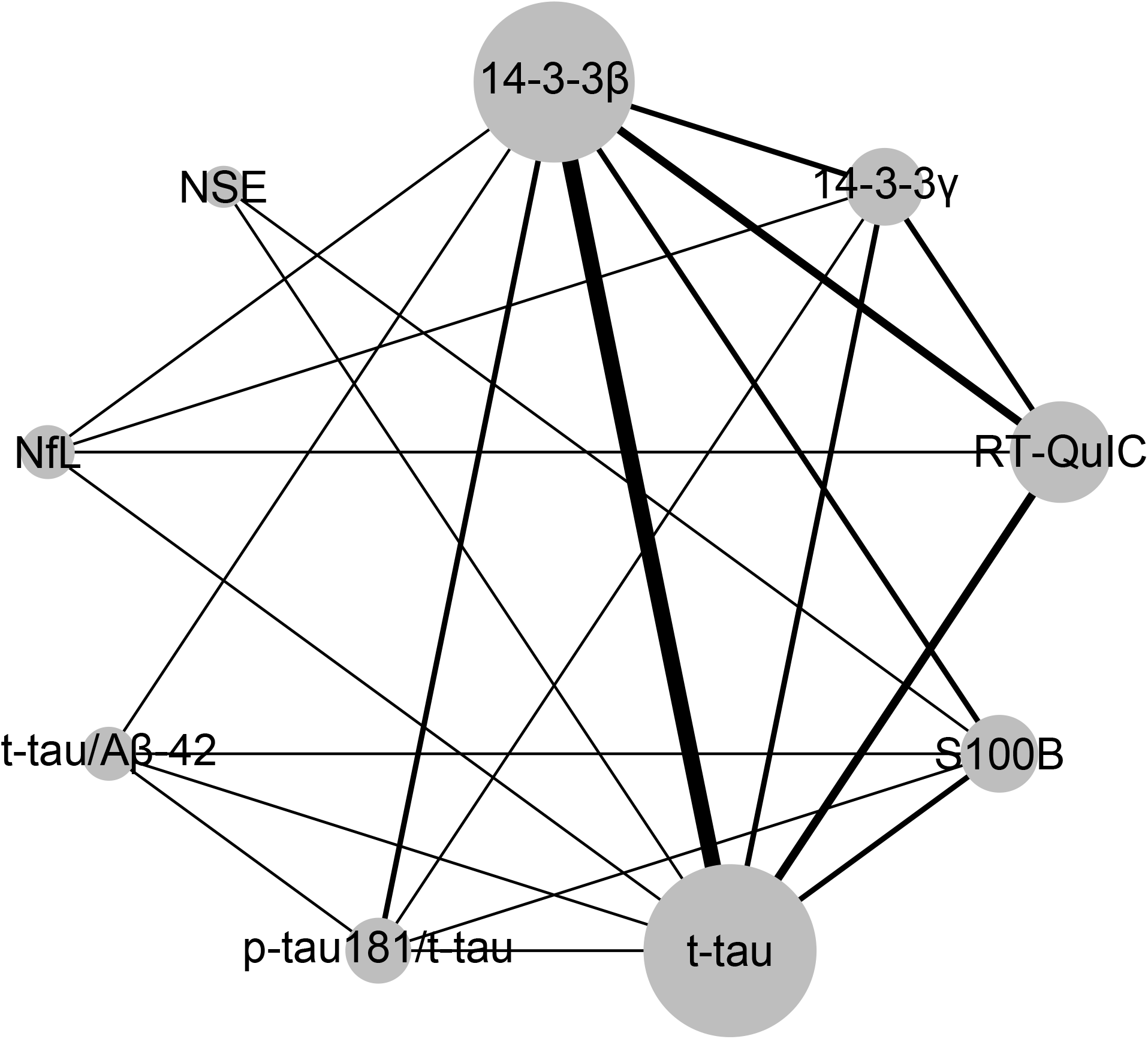
Network plot *The thickness of the nodes is proportional to the number of direct comparisons.*

**Figure 4.**
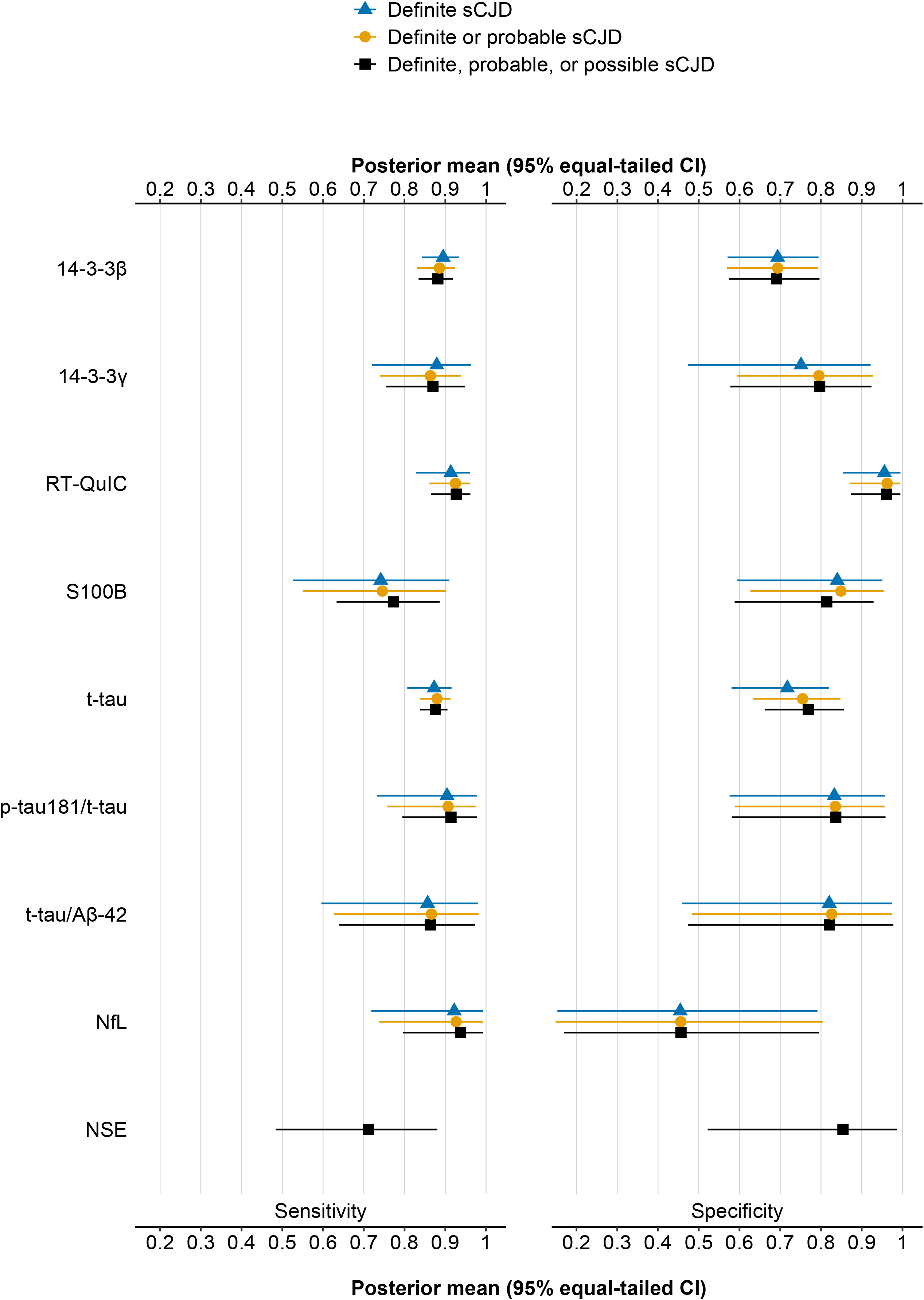
Results of network meta-analyses (stratified by level of certainty of sCJD diagnosis) *CI: Confidence interval*

The pooled sensitivities and specificities of most included biomarkers did not change much (±0.01) when including lower levels of certainty of sCJD diagnosis (Figure 4). As an exception, specificity increased from 0.75 to 0.80 for 14-3-3γ and from 0.75 to 0.77 for t-tau, while it decreased from 0.84 to 0.81 for S100B. At the same time, the sensitivity increased from 0.74 to 0.77 for S100B. RT-QuIC was the most accurate test in among all levels of certainty of sCJD diagnosis.

## Discussion

In our systematic review, we evaluated for the first time systematically the diagnostic accuracy of proposed CSF and blood biomarkers for the differential diagnosis of sCJD in a specialised care setting. We applied the innovative concept of a diagnostic network meta-analysis to compare accuracies between different biomarkers. We found that RT-QuIC had the best-balanced accuracy and specificity throughout all scenarios.

Most of the 190 diagnostic accuracy studies retrieved in this systematic review did not meet the highest quality requirements of evidence-based medicine. All diagnostic studies evaluated were subject to methodological limitations, partly due to the rare disease situation, the lack of a uniform reference standard, and various forms of bias. Even the 16 (8.4%) studies included in the final analysis had limitations mainly based on the study population’s composition and the timing of the definition of the reference standard.

Subgroup analysis results indicate that the heterogeneity observed was mainly caused by choice of study population and less by methodological quality or clinical criteria. Specificity estimates showed the highest heterogeneity since most biomarkers used in the sCJD context are unspecific markers of neurodegeneration so that the composition of the non-sCJD group is crucial. One exception was RT-QuIC, which is the only available prion-specific biomarker so far. Although we only included diagnostic studies that reflected the real-world referral setting of a specialized care centre, the proportion of other neurodegenerative diseases in the non-CJD group might have been heterogeneous based on country-specific referral patterns or local research foci.

Many methodological constraints found in our systematic review were typical for diagnostic accuracy studies dealing with low-prevalence settings. Here, the gold standard phase III diagnostic study design with the recruitment of consecutive patients suspected of the target condition who subsequently receive the index test and the reference standard is rarely applied in practice because of the large overall sample size, the long study duration and the high costs. To ensure optimal diagnostic accuracy estimates under these preconditions, Holtman and colleagues provide a general guide for six applicable designs in different low-prevalence situations.^38^ The design’s selection best suitable should be made based on the specific constraints of the particular situation (e.g. patient selection, the invasiveness of an index test or reference standard, the target condition itself). The designs suggested by Holtman and colleagues offer reasonable approaches to improve the methodological quality and evidence for diagnostic studies in sCJD. Moreover, taking into account the recommendations of the regulatory authorities EMA^39^ and FDA^40^, the Cochrane Collaboration^41^, the QUADAS-2 tool^10^, and the STARD checklist^42^ can considerably contribute to ensuring high-quality diagnostic accuracy studies.

The inclusion/exclusion criteria and signalling questions of the QUADAS-2 tool that we applied do not only represent the prerequisites for conducting valid meta-analyses, but also reflect the patient characteristics and basic medical criteria that are essential for reliable decision-making in a real-world clinical setting. In the absence of this information in diagnostic accuracy studies, such studies provide only limited evidence to support the medical diagnosis of sCJD in everyday clinical practice. If relevant patient data are not reported and cannot be retrieved, diagnostic studies may be of little use for the reader because the applicability and generalisability of the results remain unclear.

By excluding healthy individuals and focusing on patients representing true differential diagnoses such as Alzheimer’s disease or other rapid progressive dementia, studies increase their relevance for clinical decision-making. It has to be assessed carefully if certain diagnoses, which are more prevalent than sCJD and can be diagnosed easily using a set of diagnostic criteria for this disease, can be removed from the field of clinically relevant differential diagnoses of sCJD if sCJD diagnosis is already ruled out based on these alternative criteria^43^. In such a case, it may also be reasonable to consider these diagnostic tests directly in the evaluation of potential sCJD patients. Our NMA results imply that RT-QuIC is overall the most accurate biomarker with both high sensitivity and specificity. RT-QuIC has already been proposed as an addition to 14-3-3 positivity for the composite reference standard,^44^ and an updated reference standard has been validated by the authors of the German CJD guideline.^45^ However, it is currently unclear how to best combine RT-QuIC and 14-3-3 in the setting of the complex composite reference standard. A naïve approach would be to use the easily applicable, but still highly sensitive 14-3-3 as screening test or initial index test in patients with suspected sCJD. Only patients tested positively could then be further examined according to the composite reference standard, including RT-QuIC instead of 14-3-3 to confirm sCJD diagnosis. Abu-Rumeileh et al.^23^ explored the combinations of two biomarkers (a surrogate marker plus RT-QuIC). Integrating their data in our NMA found that NfL had the highest sensitivity for all certainty levels of sCJD diagnosis, but the lowest specificity, resulting in the poorest balanced accuracy. From our perspective, the issues of biomarker test combinations and screening tests need further validation. High-quality primary studies with paired information on at least two biomarkers for each individual are necessary to develop more sophisticated strategies to combine biomarkers. Moreover, estimates for the expected benefit for the patient are needed to make inferences about the likely impact on patient-important outcomes.^46^ Strengths and limitations

Our systematic review provides the most extensive evaluation of biomarkers for the differential diagnosis of sCJD. It is the first one offering head-to-head comparisons. Despite considerable heterogeneity between studies, this comparison was possible since we applied a specific form of diagnostic network meta-analysis. We only included studies that mimicked a phase III diagnostic study design, and were performed in the real-life target population. By doing so, we wanted to provide the best available evidence to support clinicians in daily real-world clinical decision-making in the differential diagnosis of sCJD. All the included studies had a moderate risk of bias in at least one of the categories assessed so that residual bias in estimates cannot be ruled out. Our systematic review included CSF and blood biomarkers, but could not identify a study based on serum or plasma biomarkers, which met our inclusion criteria. Compared to CSF biomarkers, serum biomarkers have many advantages e.g. with respect to invasiveness, patient acceptability, cost and time-effectiveness, and population-level feasibility, making them beneficial complementary partners of CSF biomarkers or neuroimaging.^47^ However, no study using serum biomarkers could be included in our systematic review. Both, comparing bio-fluid biomarkers to and combining them with neuroimaging is a relevant research field, as the recent study of Bizzi et al.^25^ showed. We included the biomarker data from this study in our NMA, but decided against an extension of our work to imaging markers, as this would result in more complex requirements about what needs to be reported in the respective studies and about how the studies were performed. This was beyond the scope of this work, but provides an interesting avenue for future studies. Due to the rare disease situation and patients’ treatment in a few specialised centres worldwide, the same patients might have been included in more than one study within the review. Although no author has explicitly described this, frozen CSF samples from one or more centres were pooled in several studies opening a potential for reuse of already analysed samples; this could have resulted in an overestimation of the study population’s variability. We did not consider that diagnostic accuracy might depend on sCJD subtype; its distribution might have affected individual study estimates. However, we only included studies that enrolled consecutive patients with suspected sCJD or a representative sample of those with suspected sCJD, so that study populations mirror the patient populations to which diagnostic tests are applied in clinical practice.

## Conclusion

Our NMA suggested RT-QuIC as the most powerful biomarker test for the differential diagnosis of sCJD. The indirect comparisons undertaken in the NMA complete the already available head-to-head evidence and confirm that RT-QuIC (potentially in combination with a more easily applicable and screening test like 14-3-3 or t-tau) should be implemented in the composite lifetime reference standard. Our work also pointed out the methodological limitations of previous diagnostic accuracy studies in the field, and the requirements we consider necessary for the design and conduct of future research projects in this setting. New high-quality studies with appropriate study designs are necessary to provide appropriate diagnostic accuracy data for unanswered research questions such as how to improve diagnostic processes in the pre-symptomatic, period and in those with atypical presentations or specific subtypes. These studies represent the evidence needed for future systematic reviews as well as (network) meta-analyses in the differential diagnosis of prion diseases.^48^

## Supporting information

PRISMA checklist

## Data Availability

All data generated or analysed during this study are included in this published article and its supplementary information files.

## Article information

## Acknowledgements

We thank all primary study authors who responded to our inquiries.

## Contributors

AK and SP designed the network meta-analysis, wrote the study protocol and conducted the systematic review (screening the literature search, selecting included studies). SP extracted clinical and demographic information, details of the assays and cut-offs used, and contacted study authors for additional information. NR and SP independently extracted the number of true and false positives and negatives. SP, AK, and NR did an independent risk of bias assessment for all included studies. NR did the statistical analysis. NR, AK and SP analysed and interpreted the data, and wrote the report’s draft with input from SK, AZ and GR. All authors critically reviewed the report and approved the final submitted manuscript version. NR and SP contributed equally as first authors. Both had full access to all of the data in the study and take responsibility for the integrity of the data and the data analysis accuracy.

