## Supplementary material for "Diagnostic accuracy of cerebrospinal fluid and blood biomarkers for the differential diagnosis of sporadic Creutzfeldt-Jakob disease: a (network) meta-analysis"

**Figure e-1: Funnel plots**

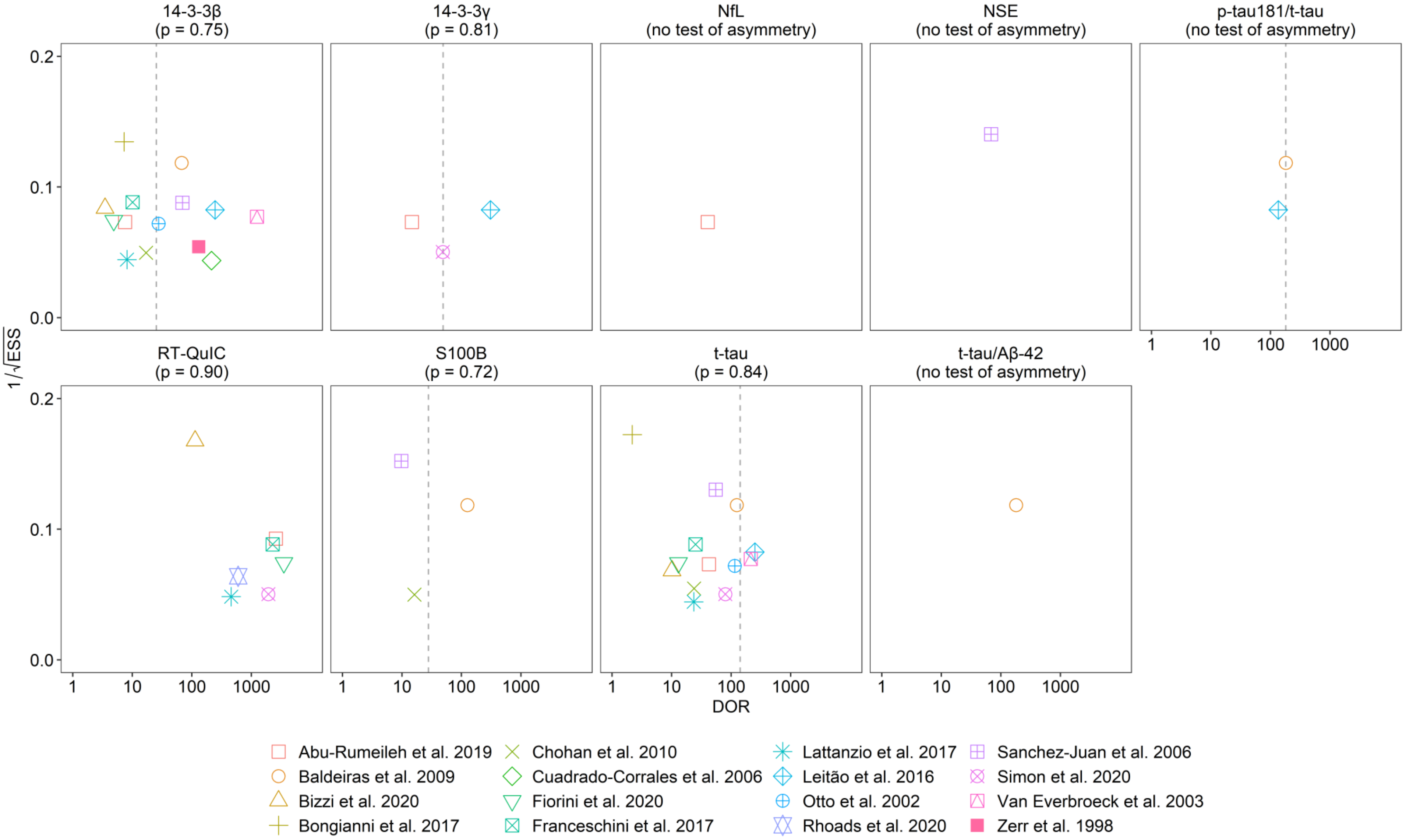

The p-value refers to the regression test of asymmetry by Deeks et al.[1]
DOR: diagnostic odds ratio
ESS: effective sample size

### Figure e-2: Sensitivity and specificity of CSF biomarkers for the diagnosis of definite, probable, or possible sCJD

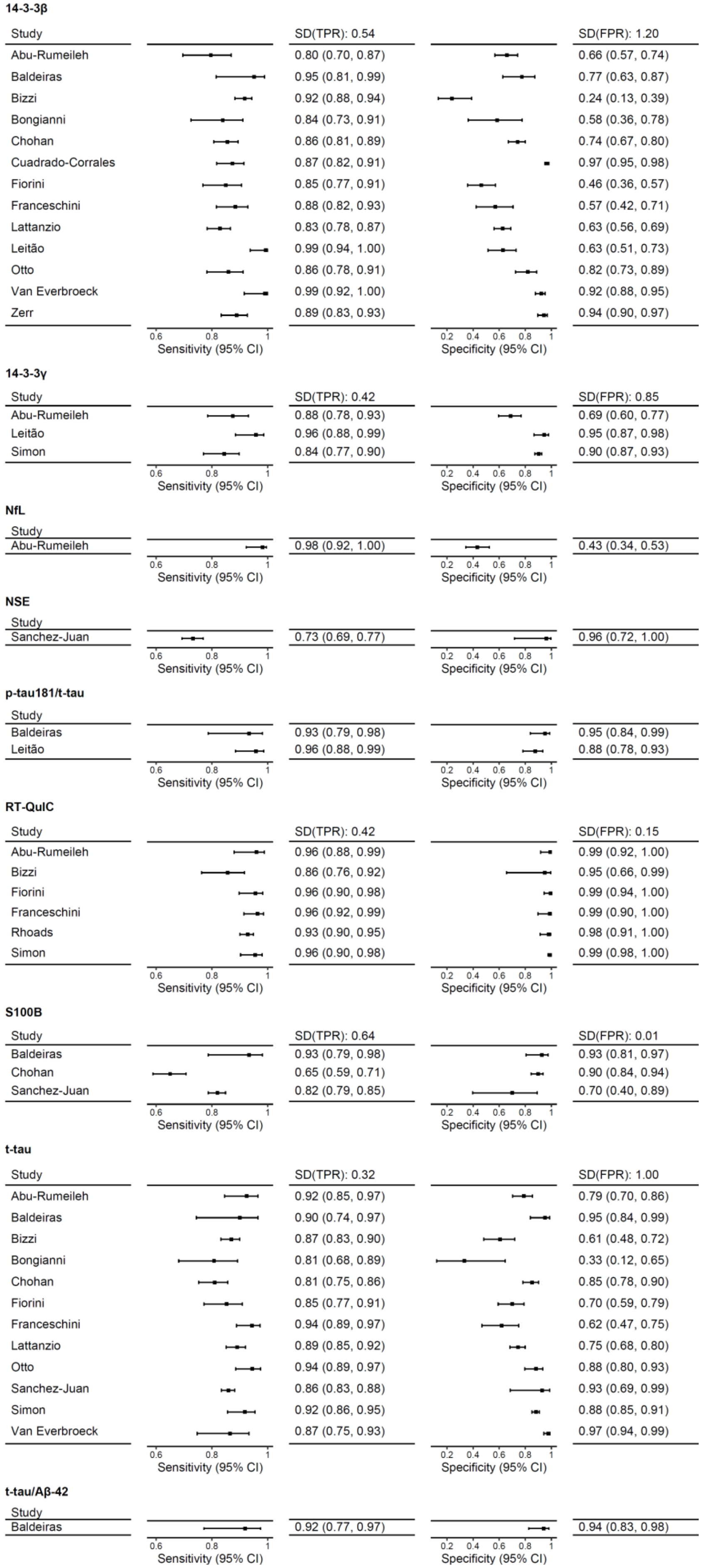

### Figure e-3: Sensitivity and specificity of CSF biomarkers for the diagnosis of definite or probable sCJD

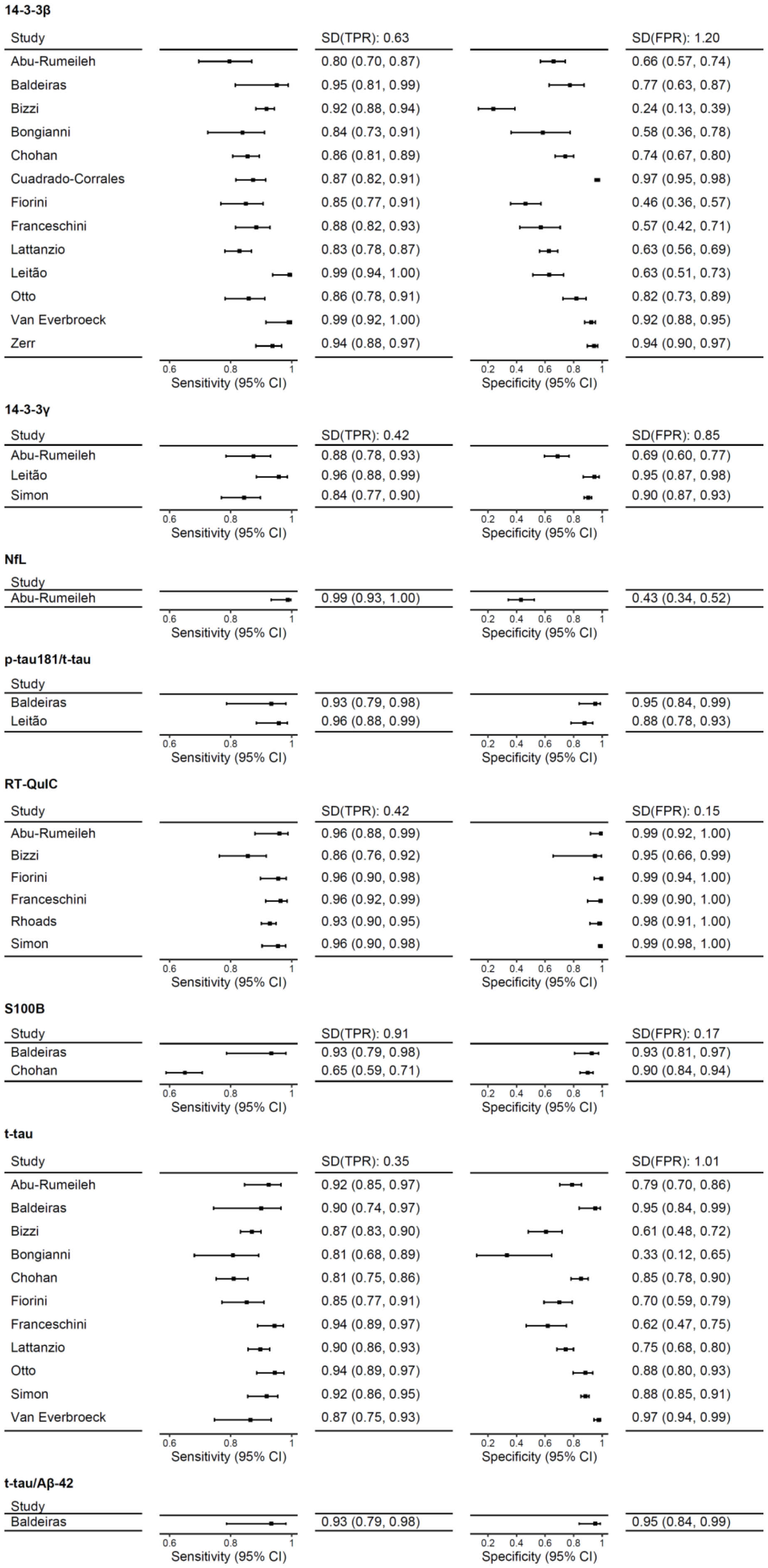

**14-3-3β**

| Study | Sensitivity (95% CI) SD(TPR): 0.63 | Specificity (95% CI) SD(FPR): 1.20 |
|---|---|---|
| Abu-Rumeileh | 0.80 (0.70, 0.87) | 0.66 (0.57, 0.74) |
| Baldeiras | 0.95 (0.81, 0.99) | 0.77 (0.63, 0.87) |
| Bizzi | 0.92 (0.88, 0.94) | 0.24 (0.13, 0.39) |
| Bongianni | 0.84 (0.73, 0.91) | 0.58 (0.36, 0.78) |
| Chohan | 0.86 (0.81, 0.89) | 0.74 (0.67, 0.80) |
| Cuadrado-Corrales | 0.87 (0.82, 0.91) | 0.97 (0.95, 0.98) |
| Fiorini | 0.85 (0.77, 0.91) | 0.46 (0.36, 0.57) |
| Franceschini | 0.88 (0.82, 0.93) | 0.57 (0.42, 0.71) |
| Lattanzio | 0.83 (0.78, 0.87) | 0.63 (0.56, 0.69) |
| Leitão | 0.99 (0.94, 1.00) | 0.63 (0.51, 0.73) |
| Otto | 0.86 (0.78, 0.91) | 0.82 (0.73, 0.89) |
| Van Everbroeck | 0.99 (0.92, 1.00) | 0.92 (0.88, 0.95) |
| Zerr | 0.94 (0.88, 0.97) | 0.94 (0.90, 0.97) |

**14-3-3γ**

| Study | Sensitivity (95% CI) SD(TPR): 0.42 | Specificity (95% CI) SD(FPR): 0.85 |
|---|---|---|
| Abu-Rumeileh | 0.88 (0.78, 0.93) | 0.69 (0.60, 0.77) |
| Leitão | 0.96 (0.88, 0.99) | 0.95 (0.87, 0.98) |
| Simon | 0.84 (0.77, 0.90) | 0.90 (0.87, 0.93) |

**NfL**

| Study | Sensitivity (95% CI) | Specificity (95% CI) |
|---|---|---|
| Abu-Rumeileh | 0.99 (0.93, 1.00) | 0.43 (0.34, 0.52) |

**p-tau181/t-tau**

| Study | Sensitivity (95% CI) | Specificity (95% CI) |
|---|---|---|
| Baldeiras | 0.93 (0.79, 0.98) | 0.95 (0.84, 0.99) |
| Leitão | 0.96 (0.88, 0.99) | 0.88 (0.78, 0.93) |

**RT-QuIC**

| Study | Sensitivity (95% CI) SD(TPR): 0.42 | Specificity (95% CI) SD(FPR): 0.15 |
|---|---|---|
| Abu-Rumeileh | 0.96 (0.88, 0.99) | 0.99 (0.92, 1.00) |
| Bizzi | 0.86 (0.76, 0.92) | 0.95 (0.66, 0.99) |
| Fiorini | 0.96 (0.90, 0.98) | 0.99 (0.94, 1.00) |
| Franceschini | 0.96 (0.92, 0.99) | 0.99 (0.90, 1.00) |
| Rhoads | 0.93 (0.90, 0.95) | 0.98 (0.91, 1.00) |
| Simon | 0.96 (0.90, 0.98) | 0.99 (0.98, 1.00) |

**S100B**

| Study | Sensitivity (95% CI) SD(TPR): 0.91 | Specificity (95% CI) SD(FPR): 0.17 |
|---|---|---|
| Baldeiras | 0.93 (0.79, 0.98) | 0.93 (0.81, 0.97) |
| Chohan | 0.65 (0.59, 0.71) | 0.90 (0.84, 0.94) |

**t-tau**

| Study | Sensitivity (95% CI) SD(TPR): 0.35 | Specificity (95% CI) SD(FPR): 1.01 |
|---|---|---|
| Abu-Rumeileh | 0.92 (0.85, 0.97) | 0.79 (0.70, 0.86) |
| Baldeiras | 0.90 (0.74, 0.97) | 0.95 (0.84, 0.99) |
| Bizzi | 0.87 (0.83, 0.90) | 0.61 (0.48, 0.72) |
| Bongianni | 0.81 (0.68, 0.89) | 0.33 (0.12, 0.65) |
| Chohan | 0.81 (0.75, 0.86) | 0.85 (0.78, 0.90) |
| Fiorini | 0.85 (0.77, 0.91) | 0.70 (0.59, 0.79) |
| Franceschini | 0.94 (0.89, 0.97) | 0.62 (0.47, 0.75) |
| Lattanzio | 0.90 (0.86, 0.93) | 0.75 (0.68, 0.80) |
| Otto | 0.94 (0.89, 0.97) | 0.88 (0.80, 0.93) |
| Simon | 0.92 (0.86, 0.95) | 0.88 (0.85, 0.91) |
| Van Everbroeck | 0.87 (0.75, 0.93) | 0.97 (0.94, 0.99) |

**t-tau/Aβ-42**

| Study | Sensitivity (95% CI) | Specificity (95% CI) |
|---|---|---|
| Baldeiras | 0.93 (0.79, 0.98) | 0.95 (0.84, 0.99) |

**Figure e-4: Heterogeneity among studies that investigated 14-3-3β**

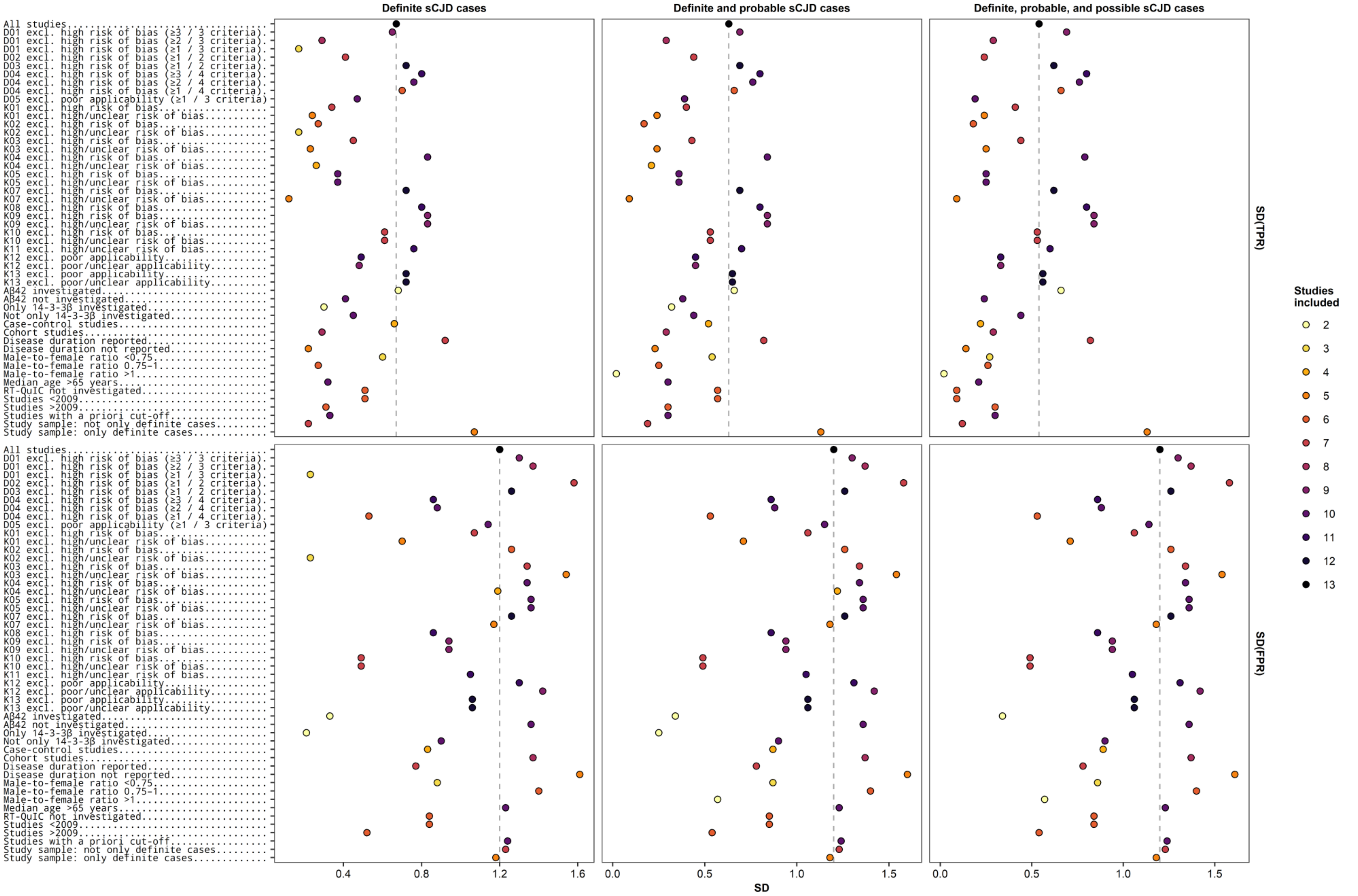

Abbreviations of domains and criteria can be found in Table e-3. FPR: false positive rate; SD: standard deviation; TPR: true positive rate

### Figure e-5: Heterogeneity among studies that investigated 14-3-3γ

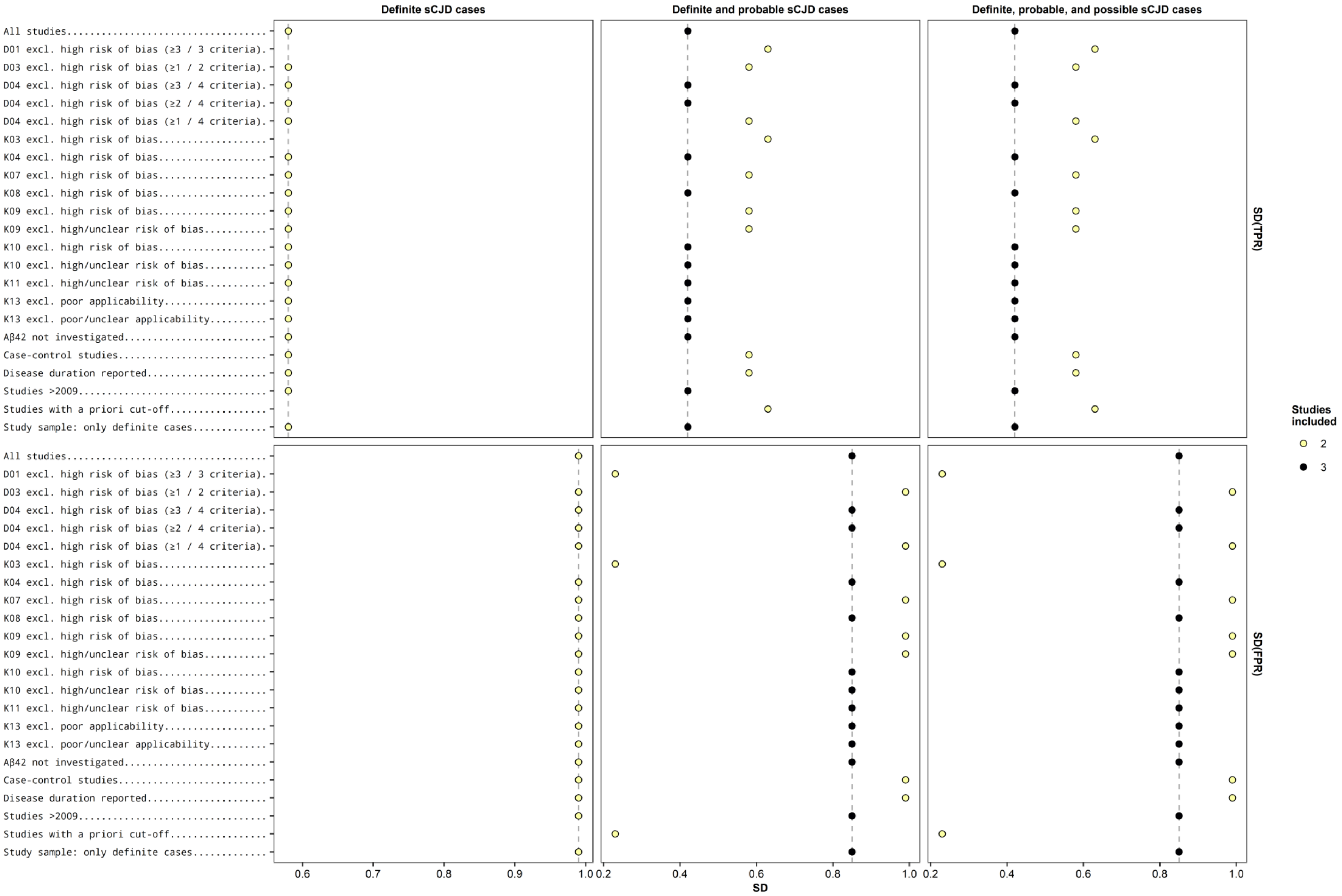

Abbreviations of domains and criteria can be found in Table e-3. FPR: false positive rate; SD: standard deviation; TPR: true positive rate

**Figure e-6: Heterogeneity among studies that investigated RT-QuIC**

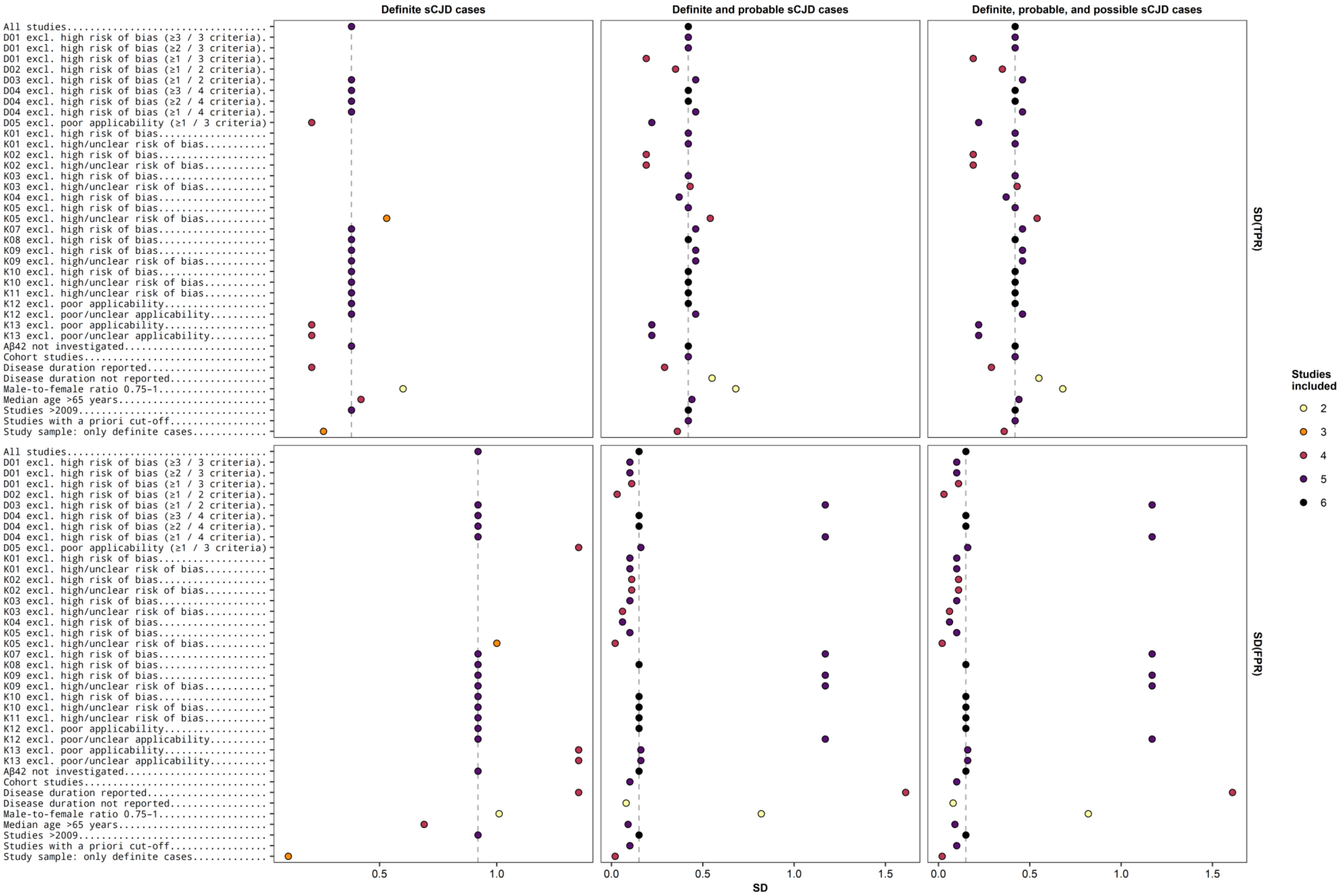

Abbreviations of domains and criteria can be found in Table e-3. FPR: false positive rate; SD: standard deviation; TPR: true positive rate

### Figure e-7: Heterogeneity among studies that investigated S100B

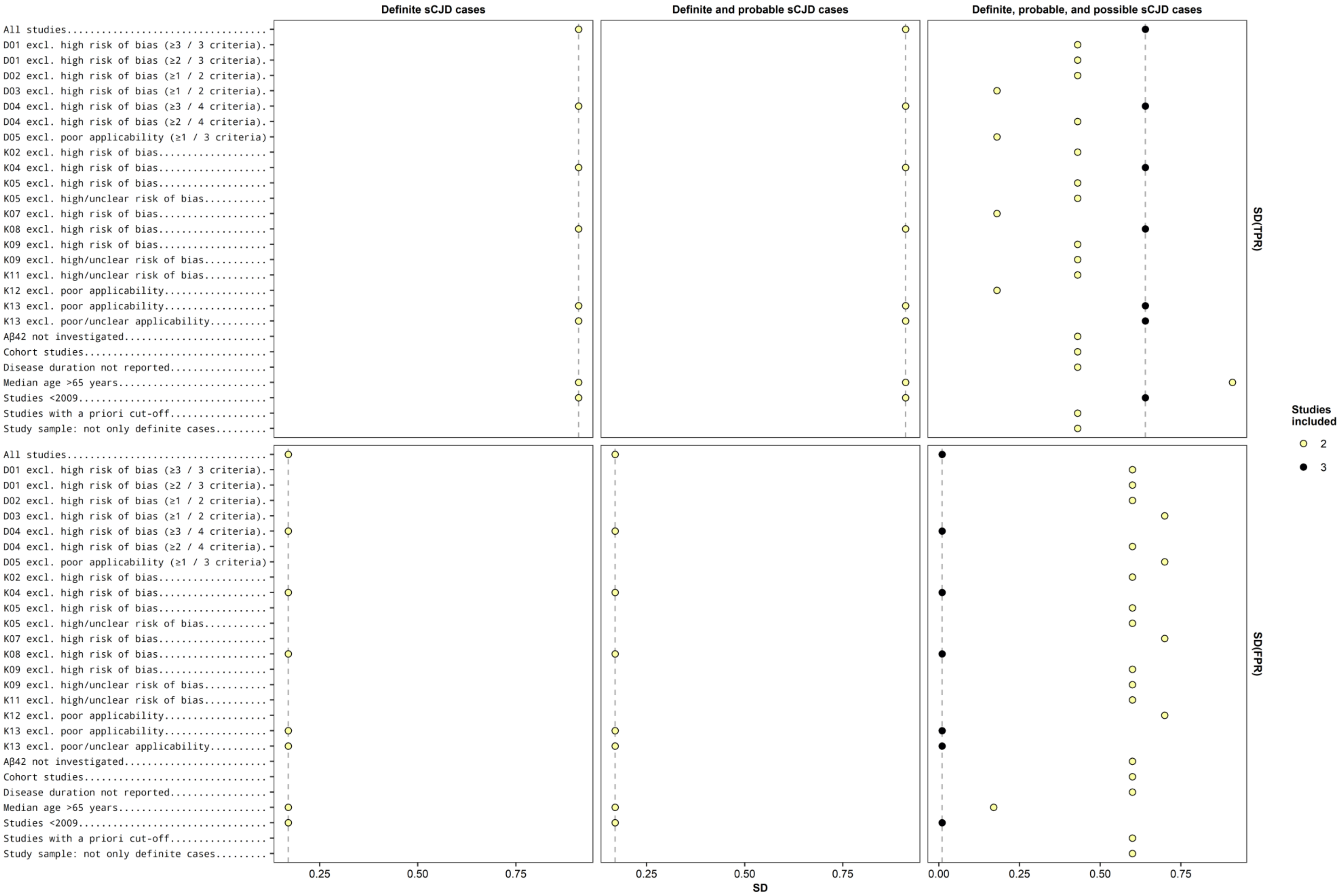

Abbreviations of domains and criteria can be found in Table e-3. FPR: false positive rate; SD: standard deviation; TPR: true positive rate

**Figure e-8: Heterogeneity among studies that investigated t-tau**

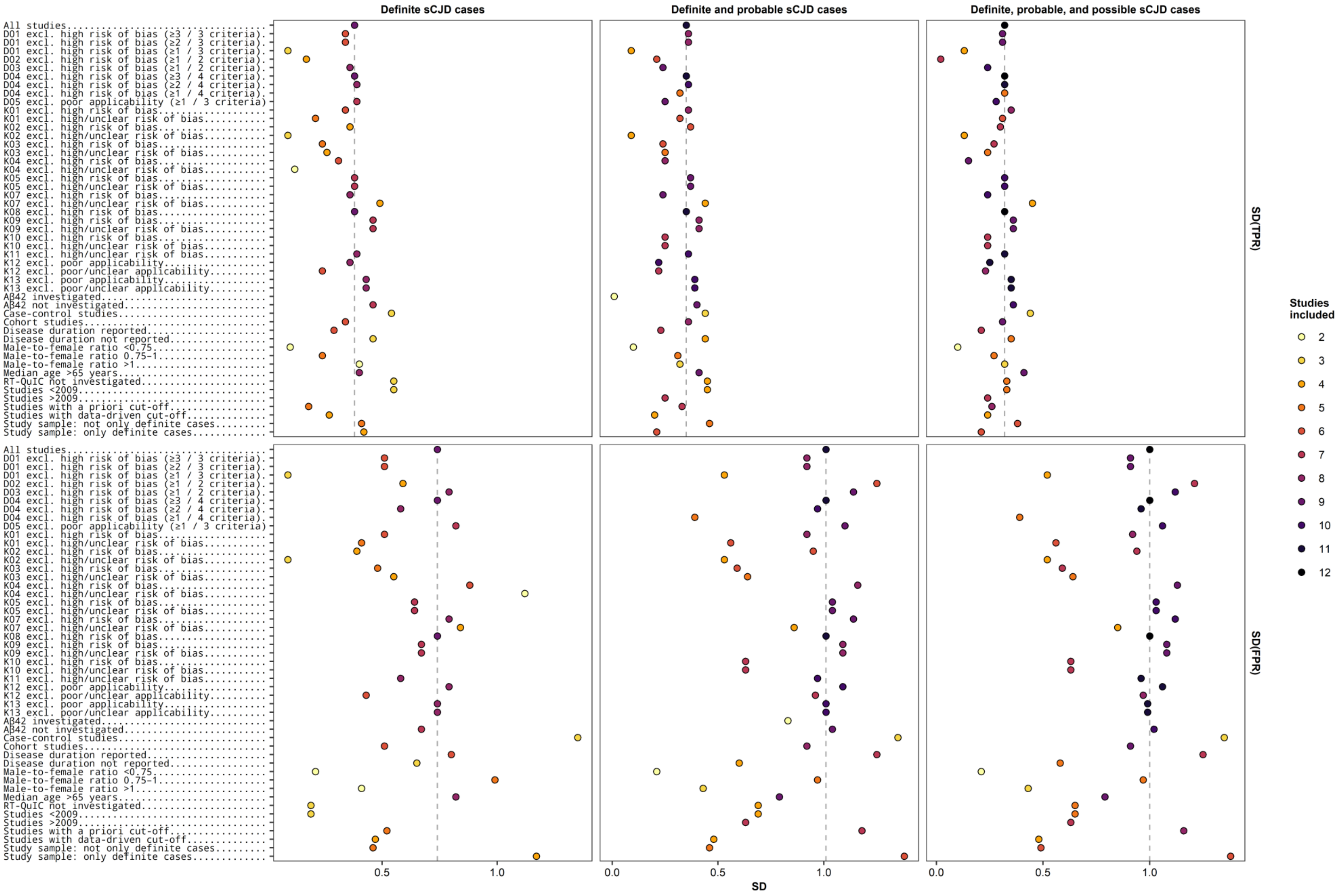

Abbreviations of domains and criteria can be found in Table e-3. FPR: false positive rate; SD: standard deviation; TPR: true positive rate

**Table e-1: Literature search strategies**

**Pubmed**

| Nr. | Query | Items found |
|---|---|---|
| #1 | "Search (((((Biomarker) OR biomarkers [MeSH Terms])) OR ((Diagnosis) OR diagnosis [MeSH Terms]))) AND ((((Creutzfeldt Jakob disease) OR CJD)) OR cjd creutzfeldt jakob disease [MeSH Terms]) AND ((CSF OR cerebrospinal) OR (blood or serum or plasma))" | 1119 |
| #2 | "Search (((((Biomarker) OR biomarkers [MeSH Terms])) OR ((Diagnosis) OR diagnosis [MeSH Terms]))) AND ((((Creutzfeldt Jakob disease) OR CJD)) OR cjd creutzfeldt jakob disease [MeSH Terms]) AND ((CSF OR cerebrospinal) OR (blood or serum or plasma))<br>Sort by: [pubsolr12]" | 1136 |

**Embase**

| Nr. | Query | Items found |
|---|---|---|
| #1 | (biomarker or diagnosis).af. | 8 865 024 |
| #2 | ((biomarker or diagnosis) and (Creutzfeldt Jakob disease or CJD) and (CSF or cerebrospinal or (blood or serum or plasma))).af. | 2651 |
| #3 | remove duplicates from 2 | 1994 |

**Cochrane Library**

| Nr. | Query | Items found |
|---|---|---|
| #1 | biomarker | 7793 |
| #2 | Any MeSH descriptor with qualifier(s): [Diagnosis - DI] | 53 513 |
| #3 | diagnosis | 12 283 |
| #4 | Creutzfeldt Jakob Disease | 89 |
| #5 | CJD | 118 |
| #6 | Any MeSH descriptor with qualifier(s): [Cerebrospinal fluid - CF] | 860 |
| #7 | cerebrospinal | 3721 |
| #8 | blood or serum or plasma | 32 966 |
| #9 | #2 or #3 | 12 283 |
| #10 | #4 or #5 | 167 |
| #11 | #6 or #7 | 3721 |
| #12 | #1 or #9 | 12 894 |
| #13 | #11 or #8 | 33 153 |
| #14 | #10 and #13 | 87 |
| #15 | #12 and #14 | 60 |

**Table e-2: Review questions and inclusion criteria**

| Category | Review question | Inclusion criterion |
|---|---|---|
| Patients | Participants with a diagnosis of a dementia subtype | Participants meet the criteria for a clinical diagnosis of any form of dementia in the specialist medical care landscape. |
| Index test | Plasma and CSF tests of biomarkers | Plasma and CSF tests of biomarkers |
| Target disease | Sporadic Creutzfeldt-Jakob disease (sCJD) | Differential diagnosis of CJD or other diseases with dementia subtypes |
| Reference standard | • ICD-10[2]<br>• Creutzfeldt-Jakob disease, 2010[3,4]<br>• Creutzfeldt-Jakob disease, stage of development: S1 (Completely revised: August 2018; Valid until: August 2023; AWMF registration number: 030/042)<br>• case definitions of the RKI for the sCJD (status 01.01.2015) | • Reference standard as stated; for differential diagnoses:<br>• NINCDS-ADRDA[5]<br>• Consortium to Establish a Registry for Alzheimer's Disease (CERAD)<br>• Alzheimer's Disease Diagnostic and Treatment Centers (ADDTC)<br>• International Classification of Diseases (ICD10)<br>• Diagnostic and Statistical Manual of Mental Disorders, Fourth Edition (DSMIV)<br>• Definitionen für die Demenz der Alzheimer Krankheit<br>• National Institute of Neurological Disorders and Stroke and Association Internationale pour la Recherché et l'Enseignement en Neurosciences (NINDS-AIREN)<br>• Cambridge Mental Disorders of the Elderly Examination (CAMDEX)<br>• Criteria acceptable for VD<br>• Lund criteria for FTD[6]<br>• BRAAK stage and McKeith criteria for DLB<br>• DSMIV for alcohol-induced dementia |
| Outcome | Not specified | Data for filling the 2x2 contingency table |
| Study design | Not specified | Cross-sectional studies in which<br>1) plasma and CSF biomarker measurements and clinical diagnostic criteria were generated in a narrow time window, and<br>2) patients with sCJD were differentiated from patients with other dementia subtypes. |

**Table e-3: QUADAS-2 items and guidelines for scoring**

| Question | Response | Explanation |
|---|---|---|
| **D01: Patient selection** | | |
| **K01: Sampling** Was the sampling method adequate? | No = high risk of bias Yes = low risk of bias | Where a sample is used, the designs with consecutive or random samples are least susceptible to producing bias. If the sample selection is based on volunteers or selected participants from a clinic or research institution, a bias is obvious. |
| **K02: Study design** Was a case-control or comparable design avoided? | No = high risk of bias Yes = low risk of bias | Design similar to the case-control approach, which could evoke bias, are those designs where the study team deliberately increases or decreases the proportion of study participants with the target disease that can no longer be representative. Some case-control methods can be excluded from the outset if they mix participants from different settings. |
| **K03: Exclusion criteria** Are the exclusion criteria described and appropriate? | No = high risk of bias Yes = low risk of bias | The study is automatically classified as unclear if the exclusions are not detailed (depending on the contact with the study authors). Where a detailed description is provided, the study will be considered low-risk if the reviewers consider the exclusion criteria appropriate. |
| **D02: Index test** | | |
| **K04: Blinding** Was the evaluation and interpretation of the biomarker tests performed without knowledge of the clinical CJD diagnosis? | No = high risk of bias Yes = low risk of bias | Terms such as "blinded" or "independent and without knowledge of" are sufficient and comprehensive details of the blinding procedure are not required. The interpretation of the results of the index test could be influenced by the knowledge of the results of the reference standard. If the index test is always evaluated before the reference standard, the evaluator may not be aware of the results of the reference standard. Then this point can be answered with "yes". For specific index tests, the result is objective, and knowledge of the reference standard should not influence the results, e.g. for protein levels in cerebrospinal fluid the quality rating, in this case, is "low risk" even if the blinding has not been implemented. |
| **K05: A priori cutoff** Were the biomarker limits pre-specified? | No = high risk of bias Yes = low risk of bias | For scales and biomarkers, there is often a reference point (in units or categories) above which individuals are classified as "test positive". This can be associated with a limit value, a clinical cut-off or a dichotomization point. A study is considered to have a high bias risk once the authors have determined the optimal cut-off post hoc from their study data, as the selection of the threshold to maximize sensitivity and specificity may lead to an over-optimistic estimate of test performance. Some publications may use an alternative method of analysis without applying a limit. These publications should be classified as "not applicable". |
| **D03: Reference standard** | | |
| **K06: Clinical diagnosis** Is the evaluation used for the clinical diagnosis of sCJD acceptable? | No = high risk of bias Yes = low risk of bias | Generally accepted international criteria to support the diagnosis of dementia are explained in ICD-10 and DSM-IV. Criteria for clinical diagnosis of sCJD can also be found in ICD-10[2] and CJD 2010[3,4],. Specific criteria for dementia subtypes include but are not limited to the NINCDSADRDA criteria for Alzheimer's disease; the McKeith criteria for Lewy body dementia; the Lund criteria for frontotemporal dementia; and the NINDS-AIREN criteria for vascular dementia. If the criteria applied are not familiar ("unclear") to the review authors, this point should be assessed as "high bias risk". |
| **K07: Blinding** Was the clinical evaluation of CJD performed without knowledge of the biomarkers? | No = high risk of bias Yes = low risk of bias | Terms such as "blinded" or "independent and without knowledge of" are sufficient and comprehensive details of the blinding procedure are not required. The interpretation of the results of the reference standard could be influenced by the knowledge of the results of the index test. |

| Question | Response | Explanation |
|---|---|---|
| **D04: Flow and timing** | | |
| **K08: Time interval**<br>Was there an appropriate interval between biomarker use and clinical evaluation? | No = high risk of bias<br>Yes = low risk of bias | The time interval between index test and reference standard influences the test accuracy. A time variable is therefore used in the evaluation. Moreover, its influence on the test accuracy is investigated. The minimum interval for a follow-up evaluation is set to 1 year. If more than 16% of the participants have received an examination, this aspect is assessed as "no". |
| **K09: Equal treatment**<br>Did all patients undergo the same evaluation for dementia, regardless of the biomarkers? | No = high risk of bias<br>Yes = low risk of bias | There may be scenarios in which people tested positive with the index test receive a more detailed examination. Where the assessment of dementia varies between individuals, the study should be rated with a high risk of bias. |
| **K10: Final analysis**<br>Were all patients who received a biomarker study included in the final analysis? | No = high risk of bias<br>Yes = low risk of bias | If the number of patients included differs from those shown in the 2x2 contingency table, distortions might occur: If the patients missing due to drop-outs differ systematically from the remaining patients, the estimators of the test performance may differ. If drop-outs were present, they should be quantified. A maximum of 20% has proven to be the maximum proportion to guarantee a low risk of distortion. |
| **K11: Missing values**<br>Have missing or non-interprTable e-biomarker test results been reported? | No = high risk of bias<br>Yes = low risk of bias | For reports of missing or non-interprTable e-values where there is a significant decrease (arbitrary value of 50% missing data), this should be classified as "no".<br>If such results have not been reported, this should be considered "unclear", and the authors should be contacted. |
| **D05: Applicability** | | |
| **K12: Representativity**<br>Were the patients included representative of the general target population? | | The included patients should be consistent with the target population described in the research question of the review. The characterization of the population is done in the form of symptoms, pre-tests, potential disease prevalence, and setting. If there are clear reasons for suspecting an unrepresentative spectrum, this aspect should be considered as little applicable. |
| **K13: Repeatability**<br>Were reliable biomarker application data available for the tests to be repeated in independent studies? | | Variations in technology, test performance, and interpretation may affect the estimation of accuracy. Also, the background and training/expertise of the assessor should be reported. If the plasma and CSF biomarkers were not consistently applied, this aspect should be assessed as poorly applicable. |
| **K14: Recency**<br>Has the clinical diagnosis of sCJD been made in a manner consistent with current clinical practice? | | For many reviews, the inclusion criteria and bias risk assessment of a CJD diagnosis will have already been assessed. For some reviews, a judgement on the applicability of the reference standard may not be available. There is a possibility that some form of CJD assessment, although valid, may lead to a diagnosis in a much larger proportion of individuals with the disease than in routine clinical practice. In this case, the aspect should be assessed as poorly applicable. |

**Table e-4: Consensus QUADAS-2 assessment and inter-rater reliability**

| | Patient selection | | | Index test | | Reference standard | | Flow and timing | | | | Applicability | | |
|---|---|---|---|---|---|---|---|---|---|---|---|---|---|---|
| | Sampling | Study design | Exclusion criteria | Blinding | A priori cutoff | Clinical diagnosis | Blinding | Time interval | Equal treatment | Final analysis | Missing values | Represen-tativity | Repeatability | Recency |
| Abu-Rumeileh et al. 2019[7] | red | red | red | yellow | red | green | yellow | yellow | green | green | green | green | green | green |
| Baldeiras et al. 2009[8] | red | red | red | yellow | red | green | yellow | yellow | red | red | yellow | yellow | green | green |
| Bizzi et al. 2020[9] | green | red | green | yellow | green | green | yellow | yellow | green | green | green | green | red | green |
| Bongianni et al. 2017[10] | red | red | red | green | green | green | green | green | green | green | green | yellow † | green | green |
| Chohan et al. 2010[11] | green | yellow | red | green | green | green | red | yellow | green | red | green | red | green | green |
| Cuadrado-Corrales et al. 2006[12] | red | yellow | green | green | green | green | green | red | red | red | yellow | green | green | green |
| Fiorini et al. 2020[13] | green | green | yellow | yellow | green | green | yellow | yellow | green | green | green | green | green | green |
| Franceschini et al. 2017[14] | green | green | green | red | green | green | green | yellow | green | green | green | green | green | green |
| (Hamlin et al. 2012[15]) | red | red | red | yellow | red | red | red | red | red | green | yellow | red | red | red |
| Lattanzio et al. 2017[16] | green | green | green | red * | green | green | green | yellow | red | green | green | green | green | green |
| Leitão et al. 2016[17] | red | red | yellow | yellow | red | green | yellow | yellow | green | green | green | red | green | green |
| Otto et al. 2002[18] | yellow | red | green | red | green | green | green | yellow | green | red | green | green | green | green |
| Rhoads et al. 2020[19] | green | green | green | green | yellow | green | yellow | yellow | green | green | green | green | green | green |
| (Rudge et al. 2018[20]) | red | red | yellow | red | yellow | yellow | yellow | yellow | yellow | yellow | yellow | red | red | red |
| Sanchez-Juan et al. 2006[21] | red | yellow | yellow | yellow | green | green | yellow | yellow | green | red | green | green | green | green |
| Simon et al. 2020[22] | green | green | green | yellow | green | green | red | yellow | red | green | green | yellow | green | green |
| Van Everbroeck et al. 2003[23] | yellow | yellow | red | yellow | green | green | green | green | yellow | green | yellow | green | green | green |
| (Wang et al. 2013[24]) | red | red | yellow | yellow | green | green | yellow | yellow | yellow | red | yellow | red | red | red |
| Zerr et al. 1998[25] | red | red | red | green | green | green | yellow | red | red | red | green | green | green | green |
| Krippendorff's alpha | 0.20 | 0.28 | 0.77 | 0.33 | 0.51 | -0.04 | 0.26 | 0.20 | 0.51 | 0.41 | 0.22 | -0.24 | -0.12 | -0.08 |

(red) = high risk of bias / poor applicability
(yellow) = unclear risk of bias / limited applicability
(green) = low risk of bias / good applicability
(red) * = low and high risk of bias next to each other (high risk of bias used in further analyses)
(yellow) † = good and limited applicability next to each other (limited applicability used in further analyses)
(X) = not included in quantitative analyses
X (blue) = authors answered inquiries

**Table e-5: Results of meta-analyses involving 14-3-3β and subgroup analyses based on QUADAS-2 quality and clinical criteria**

Abbreviations of domains and criteria can be found in Table e-3.
FPR: false positive rate
NaN: not a number (correlation cannot be calculated)
SD: standard deviation
TPR: true positive rate

| | | | | | **Studies that investigated 14-3-3β (×: included in respective analysis)** | | | | | | | | | | | | |
|---|---|---|---|---|---|---|---|---|---|---|---|---|---|---|---|---|---|
| **Level of certainty of sCJD diagnosis** | **Studies included** | **SD (TPR)** | **SD (FPR)** | **Corr.** | **Abu-Rumeileh** | **Baldeiras** | **Bizzi** | **Bongianni** | **Chohan** | **Cuadrado-Corrales** | **Fiorini** | **Franceschini** | **Lattanzio** | **Leitão** | **Otto** | **Van Everbroeck** | **Zerr** |
| **All studies** | | | | | | | | | | | | | | | | | |
| Definite, probable, and possible sCJD cases | 13 | 0.54 | 1.20 | 0.11 | × | × | × | × | × | × | × | × | × | × | × | × | × |
| Definite and probable sCJD cases | 13 | 0.63 | 1.20 | 0.23 | × | × | × | × | × | × | × | × | × | × | × | × | × |
| Definite sCJD cases | 13 | 0.67 | 1.20 | 0.24 | × | × | × | × | × | × | × | × | × | × | × | × | × |
| **Aβ42 investigated** | | | | | | | | | | | | | | | | | |
| Definite, probable, and possible sCJD cases | 2 | 0.66 | 0.34 | 1.00 | | × | | | | | | | × | | | | |
| Definite and probable sCJD cases | 2 | 0.66 | 0.34 | 1.00 | | × | | | | | | | × | | | | |
| Definite sCJD cases | 2 | 0.68 | 0.33 | 1.00 | | × | | | | | | | × | | | | |
| **Aβ42 not investigated** | | | | | | | | | | | | | | | | | |
| Definite, probable, and possible sCJD cases | 10 | 0.24 | 1.36 | 0.06 | × | | × | × | × | × | × | × | | | × | × | × |
| Definite and probable sCJD cases | 10 | 0.38 | 1.36 | 0.31 | × | | × | × | × | × | × | × | | | × | × | × |
| Definite sCJD cases | 10 | 0.41 | 1.36 | 0.31 | × | | × | × | × | × | × | × | | | × | × | × |
| **Case-control studies** | | | | | | | | | | | | | | | | | |
| Definite, probable, and possible sCJD cases | 4 | 0.22 | 0.89 | 1.00 | × | × | | × | | | | | | | | | × |
| Definite and probable sCJD cases | 4 | 0.52 | 0.87 | 1.00 | × | × | | × | | | | | | | | | × |
| Definite sCJD cases | 4 | 0.66 | 0.83 | 1.00 | × | × | | × | | | | | | | | | × |
| **Cohort studies** | | | | | | | | | | | | | | | | | |
| Definite, probable, and possible sCJD cases | 8 | 0.29 | 1.37 | 0.05 | | | × | | × | × | × | × | × | | × | × | |
| Definite and probable sCJD cases | 8 | 0.29 | 1.37 | 0.05 | | | × | | × | × | × | × | × | | × | × | |
| Definite sCJD cases | 8 | 0.29 | 1.37 | 0.08 | | | × | | × | × | × | × | × | | × | × | |
| **D01 excl. high risk of bias (≥3 / 3 criteria)** | | | | | | | | | | | | | | | | | |
| Definite, probable, and possible sCJD cases | 9 | 0.69 | 1.30 | 0.10 | | | × | | × | × | × | × | × | × | × | × | |
| Definite and probable sCJD cases | 9 | 0.69 | 1.30 | 0.10 | | | × | | × | × | × | × | × | × | × | × | |

| Level of certainty of sCJD diagnosis | Studies included | SD (TPR) | SD (FPR) | Corr. | Studies that investigated 14-3-3β (×: included in respective analysis) | | | | | | | | | | | | |
|---|---|---|---|---|---|---|---|---|---|---|---|---|---|---|---|---|---|
| | | | | | Abu-Rumeileh | Baldeiras | Bizzi | Bongianni | Chohan | Cuadrado-Corrales | Fiorini | Franceschini | Lattanzio | Leitão | Otto | Van Everbroeck | Zerr |
| Definite sCJD cases | 9 | 0.65 | 1.30 | 0.12 | | | × | | × | × | × | × | × | × | × | × | |
| **D01 excl. high risk of bias (≥2 / 3 criteria)** | | | | | | | | | | | | | | | | | |
| Definite, probable, and possible sCJD cases | 8 | 0.29 | 1.37 | 0.05 | | | × | | × | × | × | × | × | | × | × | |
| Definite and probable sCJD cases | 8 | 0.29 | 1.37 | 0.05 | | | × | | × | × | × | × | × | | × | × | |
| Definite sCJD cases | 8 | 0.29 | 1.37 | 0.08 | | | × | | × | × | × | × | × | | × | × | |
| **D01 excl. high risk of bias (≥1 / 3 criteria)** | | | | | | | | | | | | | | | | | |
| Definite, probable, and possible sCJD cases | 3 | 0 | 0.22 | NaN | | | | | | | × | × | × | | | | |
| Definite and probable sCJD cases | 3 | 0 | 0.22 | NaN | | | | | | | × | × | × | | | | |
| Definite sCJD cases | 3 | 0.17 | 0.23 | -1.00 | | | | | | | × | × | × | | | | |
| **D02 excl. high risk of bias (≥1 / 2 criteria)** | | | | | | | | | | | | | | | | | |
| Definite, probable, and possible sCJD cases | 7 | 0.24 | 1.58 | 0.01 | | | × | × | × | × | × | | | | | × | × |
| Definite and probable sCJD cases | 7 | 0.44 | 1.58 | 0.32 | | | × | × | × | × | × | | | | | × | × |
| Definite sCJD cases | 7 | 0.41 | 1.58 | 0.34 | | | × | × | × | × | × | | | | | × | × |
| **D03 excl. high risk of bias (≥1 / 2 criteria)** | | | | | | | | | | | | | | | | | |
| Definite, probable, and possible sCJD cases | 12 | 0.62 | 1.26 | 0.12 | × | × | × | × | | × | × | × | × | × | × | × | × |
| Definite and probable sCJD cases | 12 | 0.69 | 1.26 | 0.24 | × | × | × | × | | × | × | × | × | × | × | × | × |
| Definite sCJD cases | 12 | 0.72 | 1.26 | 0.25 | × | × | × | × | | × | × | × | × | × | × | × | × |
| **D04 excl. high risk of bias (≥3 / 4 criteria)** | | | | | | | | | | | | | | | | | |
| Definite, probable, and possible sCJD cases | 11 | 0.80 | 0.86 | 0.37 | × | × | × | × | × | | × | × | × | × | × | × | |
| Definite and probable sCJD cases | 11 | 0.80 | 0.86 | 0.37 | × | × | × | × | × | | × | × | × | × | × | × | |
| Definite sCJD cases | 11 | 0.80 | 0.86 | 0.30 | × | × | × | × | × | | × | × | × | × | × | × | |
| **D04 excl. high risk of bias (≥2 / 4 criteria)** | | | | | | | | | | | | | | | | | |
| Definite, probable, and possible sCJD cases | 10 | 0.76 | 0.88 | 0.32 | × | | × | × | × | | × | × | × | × | × | × | |
| Definite and probable sCJD cases | 10 | 0.76 | 0.88 | 0.32 | × | | × | × | × | | × | × | × | × | × | × | |
| Definite sCJD cases | 10 | 0.76 | 0.88 | 0.25 | × | | × | × | × | | × | × | × | × | × | × | |
| **D04 excl. high risk of bias (≥1 / 4 criteria)** | | | | | | | | | | | | | | | | | |
| Definite, probable, and possible sCJD cases | 6 | 0.66 | 0.53 | -0.08 | × | | × | × | | | × | × | | × | | | |
| Definite and probable sCJD cases | 6 | 0.66 | 0.53 | -0.08 | × | | × | × | | | × | × | | × | | | |
| Definite sCJD cases | 6 | 0.70 | 0.53 | -0.01 | × | | × | × | | | × | × | | × | | | |

| Level of certainty of sCJD diagnosis | Studies included | SD (TPR) | SD (FPR) | Corr. | Abu-Rumeileh | Baldeiras | Bizzi | Bongianni | Chohan | Cuadrado-Corrales | Fiorini | Franceschini | Lattanzio | Leitão | Otto | Van Everbroeck | Zerr |
|---|---|---|---|---|---|---|---|---|---|---|---|---|---|---|---|---|---|
| | | | | | Studies that investigated 14-3-3β (×: included in respective analysis) | | | | | | | | | | | | |
| **D05 excl. poor applicability (≥1 / 3 criteria)** | | | | | | | | | | | | | | | | | |
| Definite, probable, and possible sCJD cases | 10 | 0.19 | 1.14 | 0.95 | × | × | | × | | × | × | × | × | | × | × | × |
| Definite and probable sCJD cases | 10 | 0.39 | 1.15 | 0.70 | × | × | | × | | × | × | × | × | | × | × | × |
| Definite sCJD cases | 10 | 0.47 | 1.14 | 0.70 | × | × | | × | | × | × | × | × | | × | × | × |
| **Disease duration not reported** | | | | | | | | | | | | | | | | | |
| Definite, probable, and possible sCJD cases | 5 | 0.14 | 1.61 | -1.00 | | | × | | × | × | | | | | × | | × |
| Definite and probable sCJD cases | 5 | 0.23 | 1.60 | -0.29 | | | × | | × | × | | | | | × | | × |
| Definite sCJD cases | 5 | 0.22 | 1.61 | -0.24 | | | × | | × | × | | | | | × | | × |
| **Disease duration reported** | | | | | | | | | | | | | | | | | |
| Definite, probable, and possible sCJD cases | 7 | 0.82 | 0.78 | 0.94 | × | × | | × | | | × | × | × | | | × | |
| Definite and probable sCJD cases | 7 | 0.82 | 0.78 | 0.94 | × | × | | × | | | × | × | × | | | × | |
| Definite sCJD cases | 7 | 0.92 | 0.77 | 0.89 | × | × | | × | | | × | × | × | | | × | |
| **K01 excl. high risk of bias** | | | | | | | | | | | | | | | | | |
| Definite, probable, and possible sCJD cases | 7 | 0.41 | 1.06 | 0.29 | | | × | | × | | × | × | × | | × | × | |
| Definite and probable sCJD cases | 7 | 0.40 | 1.06 | 0.29 | | | × | | × | | × | × | × | | × | × | |
| Definite sCJD cases | 7 | 0.34 | 1.07 | 0.12 | | | × | | × | | × | × | × | | × | × | |
| **K01 excl. high/unclear risk of bias** | | | | | | | | | | | | | | | | | |
| Definite, probable, and possible sCJD cases | 5 | 0.24 | 0.71 | -1.00 | | | × | | × | | × | × | × | | | | |
| Definite and probable sCJD cases | 5 | 0.24 | 0.71 | -1.00 | | | × | | × | | × | × | × | | | | |
| Definite sCJD cases | 5 | 0.24 | 0.70 | -1.00 | | | × | | × | | × | × | × | | | | |
| **K02 excl. high risk of bias** | | | | | | | | | | | | | | | | | |
| Definite, probable, and possible sCJD cases | 6 | 0.18 | 1.26 | 0.96 | | | | | × | × | × | × | × | | | × | |
| Definite and probable sCJD cases | 6 | 0.17 | 1.26 | 1.00 | | | | | × | × | × | × | × | | | × | |
| Definite sCJD cases | 6 | 0.27 | 1.26 | 1.00 | | | | | × | × | × | × | × | | | × | |
| **K02 excl. high/unclear risk of bias** | | | | | | | | | | | | | | | | | |
| Definite, probable, and possible sCJD cases | 3 | 0 | 0.22 | NaN | | | | | | | × | × | × | | | | |
| Definite and probable sCJD cases | 3 | 0 | 0.22 | NaN | | | | | | | × | × | × | | | | |
| Definite sCJD cases | 3 | 0.17 | 0.23 | -1.00 | | | | | | | × | × | × | | | | |
| **K03 excl. high risk of bias** | | | | | | | | | | | | | | | | | |

| Level of certainty of sCJD diagnosis | Studies included | SD (TPR) | SD (FPR) | Corr. | Studies that investigated 14-3-3β (×: included in respective analysis) | | | | | | | | | | | | |
|---|---|---|---|---|---|---|---|---|---|---|---|---|---|---|---|---|---|
| | | | | | Abu-Rumeileh | Baldeiras | Bizzi | Bongianni | Chohan | Cuadrado-Corrales | Fiorini | Franceschini | Lattanzio | Leitão | Otto | Van Everbroeck | Zerr |
| Definite, probable, and possible sCJD cases | 7 | 0.44 | 1.34 | -0.25 | | | × | | | × | × | × | × | × | × | | |
| Definite and probable sCJD cases | 7 | 0.43 | 1.34 | -0.25 | | | × | | | × | × | × | × | × | × | | |
| Definite sCJD cases | 7 | 0.45 | 1.34 | -0.22 | | | × | | | × | × | × | × | × | × | | |
| **K03 excl. high/unclear risk of bias** | | | | | | | | | | | | | | | | | |
| Definite, probable, and possible sCJD cases | 5 | 0.25 | 1.54 | -0.52 | | | × | | | × | | × | × | | × | | |
| Definite and probable sCJD cases | 5 | 0.24 | 1.54 | -0.54 | | | × | | | × | | × | × | | × | | |
| Definite sCJD cases | 5 | 0.23 | 1.54 | -0.59 | | | × | | | × | | × | × | | × | | |
| **K04 excl. high risk of bias** | | | | | | | | | | | | | | | | | |
| Definite, probable, and possible sCJD cases | 10 | 0.79 | 1.34 | 0.10 | × | × | × | × | × | × | × | | | × | | × | × |
| Definite and probable sCJD cases | 10 | 0.84 | 1.34 | 0.21 | × | × | × | × | × | × | × | | | × | | × | × |
| Definite sCJD cases | 10 | 0.83 | 1.34 | 0.22 | × | × | × | × | × | × | × | | | × | | × | × |
| **K04 excl. high/unclear risk of bias** | | | | | | | | | | | | | | | | | |
| Definite, probable, and possible sCJD cases | 4 | 0 | 1.21 | NaN | | | | × | × | × | | | | | | | × |
| Definite and probable sCJD cases | 4 | 0.21 | 1.22 | 1.00 | | | | × | × | × | | | | | | | × |
| Definite sCJD cases | 4 | 0.26 | 1.19 | 1.00 | | | | × | × | × | | | | | | | × |
| **K05 excl. high risk of bias** | | | | | | | | | | | | | | | | | |
| Definite, probable, and possible sCJD cases | 10 | 0.25 | 1.36 | 0.10 | | | × | × | × | × | × | × | × | | × | × | × |
| Definite and probable sCJD cases | 10 | 0.36 | 1.36 | 0.33 | | | × | × | × | × | × | × | × | | × | × | × |
| Definite sCJD cases | 10 | 0.37 | 1.36 | 0.33 | | | × | × | × | × | × | × | × | | × | × | × |
| **K05 excl. high/unclear risk of bias** | | | | | | | | | | | | | | | | | |
| Definite, probable, and possible sCJD cases | 10 | 0.25 | 1.36 | 0.10 | | | × | × | × | × | × | × | × | | × | × | × |
| Definite and probable sCJD cases | 10 | 0.36 | 1.36 | 0.33 | | | × | × | × | × | × | × | × | | × | × | × |
| Definite sCJD cases | 10 | 0.37 | 1.36 | 0.33 | | | × | × | × | × | × | × | × | | × | × | × |
| **K07 excl. high risk of bias** | | | | | | | | | | | | | | | | | |
| Definite, probable, and possible sCJD cases | 12 | 0.62 | 1.26 | 0.12 | × | × | × | × | | × | × | × | × | × | × | × | × |
| Definite and probable sCJD cases | 12 | 0.69 | 1.26 | 0.24 | × | × | × | × | | × | × | × | × | × | × | × | × |
| Definite sCJD cases | 12 | 0.72 | 1.26 | 0.25 | × | × | × | × | | × | × | × | × | × | × | × | × |
| **K07 excl. high/unclear risk of bias** | | | | | | | | | | | | | | | | | |
| Definite, probable, and possible sCJD cases | 5 | 0.09 | 1.18 | 1.00 | | | | × | | × | | × | × | | × | | |

**Studies that investigated 14-3-3β (×: included in respective analysis)**

| Level of certainty of sCJD diagnosis | Studies included | SD (TPR) | SD (FPR) | Corr. | Abu-Rumeileh | Baldeiras | Bizzi | Bongianni | Chohan | Cuadrado-Corrales | Fiorini | Franceschini | Lattanzio | Leitão | Otto | Van Everbroeck | Zerr |
|---|---|---|---|---|---|---|---|---|---|---|---|---|---|---|---|---|---|
| Definite and probable sCJD cases | 5 | 0.09 | 1.18 | 1.00 | | | | × | | × | | × | × | | × | | |
| Definite sCJD cases | 5 | 0.12 | 1.17 | 1.00 | | | | × | | × | | × | × | | × | | |
| **K08 excl. high risk of bias** | | | | | | | | | | | | | | | | | |
| Definite, probable, and possible sCJD cases | 11 | 0.80 | 0.86 | 0.37 | × | × | × | × | × | | × | × | × | × | × | × | |
| Definite and probable sCJD cases | 11 | 0.80 | 0.86 | 0.37 | × | × | × | × | × | | × | × | × | × | × | × | |
| Definite sCJD cases | 11 | 0.80 | 0.86 | 0.30 | × | × | × | × | × | | × | × | × | × | × | × | |
| **K08 excl. high/unclear risk of bias** | | | | | | | | | | | | | | | | | |
| Definite, probable, and possible sCJD cases | 1 | | | | | | | × | | | | | | | | | |
| Definite and probable sCJD cases | 1 | | | | | | | × | | | | | | | | | |
| Definite sCJD cases | 1 | | | | | | | × | | | | | | | | | |
| **K09 excl. high risk of bias** | | | | | | | | | | | | | | | | | |
| Definite, probable, and possible sCJD cases | 9 | 0.84 | 0.94 | 0.34 | × | | × | × | × | | × | × | | × | × | × | |
| Definite and probable sCJD cases | 9 | 0.84 | 0.94 | 0.34 | × | | × | × | × | | × | × | | × | × | × | |
| Definite sCJD cases | 9 | 0.83 | 0.94 | 0.26 | × | | × | × | × | | × | × | | × | × | × | |
| **K09 excl. high/unclear risk of bias** | | | | | | | | | | | | | | | | | |
| Definite, probable, and possible sCJD cases | 9 | 0.84 | 0.94 | 0.34 | × | | × | × | × | | × | × | | × | × | × | |
| Definite and probable sCJD cases | 9 | 0.84 | 0.94 | 0.34 | × | | × | × | × | | × | × | | × | × | × | |
| Definite sCJD cases | 9 | 0.83 | 0.94 | 0.26 | × | | × | × | × | | × | × | | × | × | × | |
| **K10 excl. high risk of bias** | | | | | | | | | | | | | | | | | |
| Definite, probable, and possible sCJD cases | 7 | 0.53 | 0.49 | -0.28 | × | | × | × | | | × | × | × | × | | | |
| Definite and probable sCJD cases | 7 | 0.53 | 0.49 | -0.28 | × | | × | × | | | × | × | × | × | | | |
| Definite sCJD cases | 7 | 0.61 | 0.49 | -0.21 | × | | × | × | | | × | × | × | × | | | |
| **K10 excl. high/unclear risk of bias** | | | | | | | | | | | | | | | | | |
| Definite, probable, and possible sCJD cases | 7 | 0.53 | 0.49 | -0.28 | × | | × | × | | | × | × | × | × | | | |
| Definite and probable sCJD cases | 7 | 0.53 | 0.49 | -0.28 | × | | × | × | | | × | × | × | × | | | |
| Definite sCJD cases | 7 | 0.61 | 0.49 | -0.21 | × | | × | × | | | × | × | × | × | | | |
| **K11 excl. high/unclear risk of bias** | | | | | | | | | | | | | | | | | |
| Definite, probable, and possible sCJD cases | 11 | 0.60 | 1.05 | 0.20 | × | | × | × | × | | × | × | × | × | × | × | × |
| Definite and probable sCJD cases | 11 | 0.70 | 1.05 | 0.39 | × | | × | × | × | | × | × | × | × | × | × | × |

| Level of certainty of sCJD diagnosis | Studies included | SD (TPR) | SD (FPR) | Corr. | Studies that investigated 14-3-3β (×: included in respective analysis) | | | | | | | | | | | | |
|---|---|---|---|---|---|---|---|---|---|---|---|---|---|---|---|---|---|
| | | | | | Abu-Rumeileh | Baldeiras | Bizzi | Bongianni | Chohan | Cuadrado-Corrales | Fiorini | Franceschini | Lattanzio | Leitão | Otto | Van Everbroeck | Zerr |
| Definite sCJD cases | 11 | 0.76 | 1.05 | 0.36 | × | | × | × | × | | × | × | × | × | × | × | × |
| **K12 excl. poor applicability** | | | | | | | | | | | | | | | | | |
| Definite, probable, and possible sCJD cases | 11 | 0.33 | 1.31 | 0.19 | × | × | × | × | | × | × | × | × | | × | × | × |
| Definite and probable sCJD cases | 11 | 0.45 | 1.31 | 0.36 | × | × | × | × | | × | × | × | × | | × | × | × |
| Definite sCJD cases | 11 | 0.49 | 1.30 | 0.37 | × | × | × | × | | × | × | × | × | | × | × | × |
| **K12 excl. poor/unclear applicability** | | | | | | | | | | | | | | | | | |
| Definite, probable, and possible sCJD cases | 9 | 0.33 | 1.42 | 0.16 | × | | × | | | × | × | × | × | | × | × | × |
| Definite and probable sCJD cases | 9 | 0.45 | 1.42 | 0.34 | × | | × | | | × | × | × | × | | × | × | × |
| Definite sCJD cases | 9 | 0.48 | 1.42 | 0.39 | × | | × | | | × | × | × | × | | × | × | × |
| **K13 excl. poor applicability** | | | | | | | | | | | | | | | | | |
| Definite, probable, and possible sCJD cases | 12 | 0.56 | 1.06 | 0.25 | × | × | | × | × | × | × | × | × | × | × | × | × |
| Definite and probable sCJD cases | 12 | 0.65 | 1.06 | 0.36 | × | × | | × | × | × | × | × | × | × | × | × | × |
| Definite sCJD cases | 12 | 0.72 | 1.06 | 0.33 | × | × | | × | × | × | × | × | × | × | × | × | × |
| **K13 excl. poor/unclear applicability** | | | | | | | | | | | | | | | | | |
| Definite, probable, and possible sCJD cases | 12 | 0.56 | 1.06 | 0.25 | × | × | | × | × | × | × | × | × | × | × | × | × |
| Definite and probable sCJD cases | 12 | 0.65 | 1.06 | 0.36 | × | × | | × | × | × | × | × | × | × | × | × | × |
| Definite sCJD cases | 12 | 0.72 | 1.06 | 0.33 | × | × | | × | × | × | × | × | × | × | × | × | × |
| **Male-to-female ratio <0.75** | | | | | | | | | | | | | | | | | |
| Definite, probable, and possible sCJD cases | 3 | 0.27 | 0.86 | 1.00 | × | | | | | | | | | | × | | × |
| Definite and probable sCJD cases | 3 | 0.54 | 0.87 | 1.00 | × | | | | | | | | | | × | | × |
| Definite sCJD cases | 3 | 0.60 | 0.88 | 1.00 | × | | | | | | | | | | × | | × |
| **Male-to-female ratio >1** | | | | | | | | | | | | | | | | | |
| Definite, probable, and possible sCJD cases | 2 | 0.02 | 0.57 | 1.00 | | | | | × | | × | | | | | | |
| Definite and probable sCJD cases | 2 | 0.02 | 0.57 | 1.00 | | | | | × | | × | | | | | | |
| Definite sCJD cases | 2 | 0 | 0.57 | NaN | | | | | × | | × | | | | | | |
| **Male-to-female ratio 0.75–1** | | | | | | | | | | | | | | | | | |
| Definite, probable, and possible sCJD cases | 6 | 0.26 | 1.40 | -0.34 | | × | × | × | | × | | × | × | | | | |
| Definite and probable sCJD cases | 6 | 0.25 | 1.40 | -0.36 | | × | × | × | | × | | × | × | | | | |
| Definite sCJD cases | 6 | 0.27 | 1.40 | -0.23 | | × | × | × | | × | | × | × | | | | |

Studies that investigated 14-3-3β (×: included in respective analysis)

| Level of certainty of sCJD diagnosis | Studies included | SD (TPR) | SD (FPR) | Corr. | Abu-Rumeileh | Baldeiras | Bizzi | Bongianni | Chohan | Cuadrado-Corrales | Fiorini | Franceschini | Lattanzio | Leitão | Otto | Van Everbroeck | Zerr |
|---|---|---|---|---|---|---|---|---|---|---|---|---|---|---|---|---|---|
| **Median age >65 years** | | | | | | | | | | | | | | | | | |
| Definite, probable, and possible sCJD cases | 10 | 0.21 | 1.23 | -0.18 | × | × | × | × | × | × | | × | × | | × | | × |
| Definite and probable sCJD cases | 10 | 0.30 | 1.23 | 0.10 | × | × | × | × | × | × | | × | × | | × | | × |
| Definite sCJD cases | 10 | 0.32 | 1.23 | 0.11 | × | × | × | × | × | × | | × | × | | × | | × |
| **Not only 14-3-3β investigated** | | | | | | | | | | | | | | | | | |
| Definite, probable, and possible sCJD cases | 10 | 0.44 | 0.90 | 0.34 | × | × | × | × | × | | × | × | × | | × | × | |
| Definite and probable sCJD cases | 10 | 0.44 | 0.90 | 0.34 | × | × | × | × | × | | × | × | × | | × | × | |
| Definite sCJD cases | 10 | 0.45 | 0.90 | 0.25 | × | × | × | × | × | | × | × | × | | × | × | |
| **Only 14-3-3β investigated** | | | | | | | | | | | | | | | | | |
| Definite, probable, and possible sCJD cases | 2 | 0 | 0.00 | NaN | | | | | | × | | | | | | | × |
| Definite and probable sCJD cases | 2 | 0.32 | 0.25 | -1.00 | | | | | | × | | | | | | | × |
| Definite sCJD cases | 2 | 0.30 | 0.21 | -1.00 | | | | | | × | | | | | | | × |
| **RT-QuIC not investigated** | | | | | | | | | | | | | | | | | |
| Definite, probable, and possible sCJD cases | 6 | 0.09 | 0.84 | 1.00 | | × | | | × | × | | | | | × | × | × |
| Definite and probable sCJD cases | 6 | 0.57 | 0.85 | 0.27 | | × | | | × | × | | | | | × | × | × |
| Definite sCJD cases | 6 | 0.51 | 0.84 | 0.57 | | × | | | × | × | | | | | × | × | × |
| **Studies <2009** | | | | | | | | | | | | | | | | | |
| Definite, probable, and possible sCJD cases | 6 | 0.09 | 0.84 | 1.00 | | × | | | × | × | | | | | × | × | × |
| Definite and probable sCJD cases | 6 | 0.57 | 0.85 | 0.27 | | × | | | × | × | | | | | × | × | × |
| Definite sCJD cases | 6 | 0.51 | 0.84 | 0.57 | | × | | | × | × | | | | | × | × | × |
| **Studies >2009** | | | | | | | | | | | | | | | | | |
| Definite, probable, and possible sCJD cases | 6 | 0.30 | 0.54 | -1.00 | × | | × | × | | | × | × | × | | | | |
| Definite and probable sCJD cases | 6 | 0.30 | 0.54 | -1.00 | × | | × | × | | | × | × | × | | | | |
| Definite sCJD cases | 6 | 0.31 | 0.52 | -1.00 | × | | × | × | | | × | × | × | | | | |
| **Studies with a priori cut-off** | | | | | | | | | | | | | | | | | |
| Definite, probable, and possible sCJD cases | 10 | 0.30 | 1.24 | 0.12 | | × | × | × | × | × | × | × | × | | × | × | |
| Definite and probable sCJD cases | 10 | 0.30 | 1.24 | 0.11 | | × | × | × | × | × | × | × | × | | × | × | |
| Definite sCJD cases | 10 | 0.33 | 1.24 | 0.13 | | × | × | × | × | × | × | × | × | | × | × | |
| **Studies with data-driven cut-off** | | | | | | | | | | | | | | | | | |

| Level of certainty of sCJD diagnosis | Studies included | SD (TPR) | SD (FPR) | Corr. | Studies that investigated 14-3-3β (×: included in respective analysis) | | | | | | | | | | | | |
|---|---|---|---|---|---|---|---|---|---|---|---|---|---|---|---|---|---|
| | | | | | Abu-Rumeileh | Baldeiras | Bizzi | Bongianni | Chohan | Cuadrado-Corrales | Fiorini | Franceschini | Lattanzio | Leitão | Otto | Van Everbroeck | Zerr |
| Definite, probable, and possible sCJD cases | 1 | | | | | | | | | | | | | | | | × |
| Definite and probable sCJD cases | 1 | | | | | | | | | | | | | | | | × |
| Definite sCJD cases | 1 | | | | | | | | | | | | | | | | × |
| **Study sample: not only definite cases** | | | | | | | | | | | | | | | | | |
| Definite, probable, and possible sCJD cases | 7 | 0.12 | 1.23 | 1.00 | | | | | × | × | × | × | × | | × | | × |
| Definite and probable sCJD cases | 7 | 0.19 | 1.23 | 1.00 | | | | | × | × | × | × | × | | × | | × |
| Definite sCJD cases | 7 | 0.22 | 1.23 | 1.00 | | | | | × | × | × | × | × | | × | | × |
| **Study sample: only definite cases** | | | | | | | | | | | | | | | | | |
| Definite, probable, and possible sCJD cases | 5 | 1.13 | 1.18 | 0.64 | × | × | × | × | | | | | | | | × | |
| Definite and probable sCJD cases | 5 | 1.13 | 1.18 | 0.64 | × | × | × | × | | | | | | | | × | |
| Definite sCJD cases | 5 | 1.07 | 1.18 | 0.62 | × | × | × | × | | | | | | | | × | |

**Table e-6: Results of meta-analyses involving 14-3-3γ and subgroup analyses based on QUADAS-2 quality and clinical criteria**
Abbreviations of domains and criteria can be found in Table e-3.
FPR: false positive rate
NaN: not a number (correlation cannot be calculated)
SD: standard deviation
TPR: true positive rate
×: included in respective analysis

| Level of certainty of sCJD diagnosis | Studies included | SD (TPR) | SD (FPR) | Corr. | Abu-Rumeileh | Leitão | Simon |
|---|---|---|---|---|---|---|---|
| **All studies** | | | | | | | |
| Definite sCJD cases | 2 | 0.58 | 0.99 | 1.00 | × | × | |
| Definite and probable sCJD cases | 3 | 0.42 | 0.85 | 0.61 | × | × | × |
| Definite, probable, and possible sCJD cases | 3 | 0.42 | 0.85 | 0.61 | × | × | × |
| **Aβ42 not investigated** | | | | | | | |
| Definite sCJD cases | 2 | 0.58 | 0.99 | 1.00 | × | × | |
| Definite and probable sCJD cases | 3 | 0.42 | 0.85 | 0.61 | × | × | × |
| Definite, probable, and possible sCJD cases | 3 | 0.42 | 0.85 | 0.61 | × | × | × |
| **Case-control studies** | | | | | | | |
| Definite sCJD cases | 2 | 0.58 | 0.99 | 1.00 | × | × | |
| Definite and probable sCJD cases | 2 | 0.58 | 0.99 | 1.00 | × | × | |
| Definite, probable, and possible sCJD cases | 2 | 0.58 | 0.99 | 1.00 | × | × | |
| **Cohort studies** | | | | | | | |
| Definite sCJD cases | 0 | | | | | | |
| Definite and probable sCJD cases | 1 | | | | | | × |
| Definite, probable, and possible sCJD cases | 1 | | | | | | × |
| **D01 excl. high risk of bias (≥3 / 3 criteria)** | | | | | | | |
| Definite sCJD cases | 1 | | | | | × | |
| Definite and probable sCJD cases | 2 | 0.63 | 0.23 | 1.00 | | × | × |
| Definite, probable, and possible sCJD cases | 2 | 0.63 | 0.23 | 1.00 | | × | × |
| **D01 excl. high risk of bias (≥2 / 3 criteria)** | | | | | | | |
| Definite sCJD cases | 0 | | | | | | |
| Definite and probable sCJD cases | 1 | | | | | | × |

| Level of certainty of sCJD diagnosis | Studies included | SD (TPR) | SD (FPR) | Corr. | Abu-Rumeileh | Leitão | Simon |
|---|---|---|---|---|---|---|---|
| Definite, probable, and possible sCJD cases | 1 | | | | | | × |
| **D01 excl. high risk of bias (≥1 / 3 criteria)** | | | | | | | |
| Definite sCJD cases | 0 | | | | | | |
| Definite and probable sCJD cases | 1 | | | | | | × |
| Definite, probable, and possible sCJD cases | 1 | | | | | | × |
| **D02 excl. high risk of bias (≥1 / 2 criteria)** | | | | | | | |
| Definite sCJD cases | 0 | | | | | | |
| Definite and probable sCJD cases | 1 | | | | | | × |
| Definite, probable, and possible sCJD cases | 1 | | | | | | × |
| **D03 excl. high risk of bias (≥1 / 2 criteria)** | | | | | | | |
| Definite sCJD cases | 2 | 0.58 | 0.99 | 1.00 | × | × | |
| Definite and probable sCJD cases | 2 | 0.58 | 0.99 | 1.00 | × | × | |
| Definite, probable, and possible sCJD cases | 2 | 0.58 | 0.99 | 1.00 | × | × | |
| **D04 excl. high risk of bias (≥3 / 4 criteria)** | | | | | | | |
| Definite sCJD cases | 2 | 0.58 | 0.99 | 1.00 | × | × | |
| Definite and probable sCJD cases | 3 | 0.42 | 0.85 | 0.61 | × | × | × |
| Definite, probable, and possible sCJD cases | 3 | 0.42 | 0.85 | 0.61 | × | × | × |
| **D04 excl. high risk of bias (≥2 / 4 criteria)** | | | | | | | |
| Definite sCJD cases | 2 | 0.58 | 0.99 | 1.00 | × | × | |
| Definite and probable sCJD cases | 3 | 0.42 | 0.85 | 0.61 | × | × | × |
| Definite, probable, and possible sCJD cases | 3 | 0.42 | 0.85 | 0.61 | × | × | × |
| **D04 excl. high risk of bias (≥1 / 4 criteria)** | | | | | | | |
| Definite sCJD cases | 2 | 0.58 | 0.99 | 1.00 | × | × | |
| Definite and probable sCJD cases | 2 | 0.58 | 0.99 | 1.00 | × | × | |
| Definite, probable, and possible sCJD cases | 2 | 0.58 | 0.99 | 1.00 | × | × | |
| **D05 excl. poor applicability (≥1 / 3 criteria)** | | | | | | | |
| Definite sCJD cases | 1 | | | | × | | |
| Definite and probable sCJD cases | 2 | 0 | 0.69 | NaN | × | | × |
| Definite, probable, and possible sCJD cases | 2 | 0 | 0.69 | NaN | × | | × |
| **Disease duration not reported** | | | | | | | |

| Level of certainty of sCJD diagnosis | Studies included | SD (TPR) | SD (FPR) | Corr. | Abu-Rumeileh | Leitão | Simon |
|---|---|---|---|---|---|---|---|
| Definite sCJD cases | 0 | | | | | | |
| Definite and probable sCJD cases | 1 | | | | | | × |
| Definite, probable, and possible sCJD cases | 1 | | | | | | × |
| **Disease duration reported** | | | | | | | |
| Definite sCJD cases | 2 | 0.58 | 0.99 | 1.00 | × | × | |
| Definite and probable sCJD cases | 2 | 0.58 | 0.99 | 1.00 | × | × | |
| Definite, probable, and possible sCJD cases | 2 | 0.58 | 0.99 | 1.00 | × | × | |
| **K01 excl. high risk of bias** | | | | | | | |
| Definite sCJD cases | 0 | | | | | | |
| Definite and probable sCJD cases | 1 | | | | | | × |
| Definite, probable, and possible sCJD cases | 1 | | | | | | × |
| **K01 excl. high/unclear risk of bias** | | | | | | | |
| Definite sCJD cases | 0 | | | | | | |
| Definite and probable sCJD cases | 1 | | | | | | × |
| Definite, probable, and possible sCJD cases | 1 | | | | | | × |
| **K02 excl. high risk of bias** | | | | | | | |
| Definite sCJD cases | 0 | | | | | | |
| Definite and probable sCJD cases | 1 | | | | | | × |
| Definite, probable, and possible sCJD cases | 1 | | | | | | × |
| **K02 excl. high/unclear risk of bias** | | | | | | | |
| Definite sCJD cases | 0 | | | | | | |
| Definite and probable sCJD cases | 1 | | | | | | × |
| Definite, probable, and possible sCJD cases | 1 | | | | | | × |
| **K03 excl. high risk of bias** | | | | | | | |
| Definite sCJD cases | 1 | | | | | × | |
| Definite and probable sCJD cases | 2 | 0.63 | 0.23 | 1.00 | | × | × |
| Definite, probable, and possible sCJD cases | 2 | 0.63 | 0.23 | 1.00 | | × | × |
| **K03 excl. high/unclear risk of bias** | | | | | | | |
| Definite sCJD cases | 0 | | | | | | |
| Definite and probable sCJD cases | 1 | | | | | | × |

| Level of certainty of sCJD diagnosis | Studies included | SD (TPR) | SD (FPR) | Corr. | Abu-Rumeileh | Leitão | Simon |
|---|---|---|---|---|---|---|---|
| Definite, probable, and possible sCJD cases | 1 | | | | | | × |
| **K04 excl. high risk of bias** | | | | | | | |
| Definite sCJD cases | 2 | 0.58 | 0.99 | 1.00 | × | × | |
| Definite and probable sCJD cases | 3 | 0.42 | 0.85 | 0.61 | × | × | × |
| Definite, probable, and possible sCJD cases | 3 | 0.42 | 0.85 | 0.61 | × | × | × |
| **K05 excl. high risk of bias** | | | | | | | |
| Definite sCJD cases | 0 | | | | | | |
| Definite and probable sCJD cases | 1 | | | | | | × |
| Definite, probable, and possible sCJD cases | 1 | | | | | | × |
| **K05 excl. high/unclear risk of bias** | | | | | | | |
| Definite sCJD cases | 0 | | | | | | |
| Definite and probable sCJD cases | 1 | | | | | | × |
| Definite, probable, and possible sCJD cases | 1 | | | | | | × |
| **K07 excl. high risk of bias** | | | | | | | |
| Definite sCJD cases | 2 | 0.58 | 0.99 | 1.00 | × | × | |
| Definite and probable sCJD cases | 2 | 0.58 | 0.99 | 1.00 | × | × | |
| Definite, probable, and possible sCJD cases | 2 | 0.58 | 0.99 | 1.00 | × | × | |
| **K08 excl. high risk of bias** | | | | | | | |
| Definite sCJD cases | 2 | 0.58 | 0.99 | 1.00 | × | × | |
| Definite and probable sCJD cases | 3 | 0.42 | 0.85 | 0.61 | × | × | × |
| Definite, probable, and possible sCJD cases | 3 | 0.42 | 0.85 | 0.61 | × | × | × |
| **K09 excl. high risk of bias** | | | | | | | |
| Definite sCJD cases | 2 | 0.58 | 0.99 | 1.00 | × | × | |
| Definite and probable sCJD cases | 2 | 0.58 | 0.99 | 1.00 | × | × | |
| Definite, probable, and possible sCJD cases | 2 | 0.58 | 0.99 | 1.00 | × | × | |
| **K09 excl. high/unclear risk of bias** | | | | | | | |
| Definite sCJD cases | 2 | 0.58 | 0.99 | 1.00 | × | × | |
| Definite and probable sCJD cases | 2 | 0.58 | 0.99 | 1.00 | × | × | |
| Definite, probable, and possible sCJD cases | 2 | 0.58 | 0.99 | 1.00 | × | × | |
| **K10 excl. high risk of bias** | | | | | | | |

| Level of certainty of sCJD diagnosis | Studies included | SD (TPR) | SD (FPR) | Corr. | Abu-Rumeileh | Leitão | Simon |
|---|---|---|---|---|---|---|---|
| Definite sCJD cases | 2 | 0.58 | 0.99 | 1.00 | × | × | |
| Definite and probable sCJD cases | 3 | 0.42 | 0.85 | 0.61 | × | × | × |
| Definite, probable, and possible sCJD cases | 3 | 0.42 | 0.85 | 0.61 | × | × | × |
| **K10 excl. high/unclear risk of bias** | | | | | | | |
| Definite sCJD cases | 2 | 0.58 | 0.99 | 1.00 | × | × | |
| Definite and probable sCJD cases | 3 | 0.42 | 0.85 | 0.61 | × | × | × |
| Definite, probable, and possible sCJD cases | 3 | 0.42 | 0.85 | 0.61 | × | × | × |
| **K11 excl. high/unclear risk of bias** | | | | | | | |
| Definite sCJD cases | 2 | 0.58 | 0.99 | 1.00 | × | × | |
| Definite and probable sCJD cases | 3 | 0.42 | 0.85 | 0.61 | × | × | × |
| Definite, probable, and possible sCJD cases | 3 | 0.42 | 0.85 | 0.61 | × | × | × |
| **K12 excl. poor applicability** | | | | | | | |
| Definite sCJD cases | 1 | | | | × | | |
| Definite and probable sCJD cases | 2 | 0 | 0.69 | NaN | × | | × |
| Definite, probable, and possible sCJD cases | 2 | 0 | 0.69 | NaN | × | | × |
| **K12 excl. poor/unclear applicability** | | | | | | | |
| Definite sCJD cases | 1 | | | | × | | |
| Definite and probable sCJD cases | 1 | | | | × | | |
| Definite, probable, and possible sCJD cases | 1 | | | | × | | |
| **K13 excl. poor applicability** | | | | | | | |
| Definite sCJD cases | 2 | 0.58 | 0.99 | 1.00 | × | × | |
| Definite and probable sCJD cases | 3 | 0.42 | 0.85 | 0.61 | × | × | × |
| Definite, probable, and possible sCJD cases | 3 | 0.42 | 0.85 | 0.61 | × | × | × |
| **K13 excl. poor/unclear applicability** | | | | | | | |
| Definite sCJD cases | 2 | 0.58 | 0.99 | 1.00 | × | × | |
| Definite and probable sCJD cases | 3 | 0.42 | 0.85 | 0.61 | × | × | × |
| Definite, probable, and possible sCJD cases | 3 | 0.42 | 0.85 | 0.61 | × | × | × |
| **Male-to-female ratio <0.75** | | | | | | | |
| Definite sCJD cases | 1 | | | | × | | |
| Definite and probable sCJD cases | 1 | | | | × | | |

| Level of certainty of sCJD diagnosis | Studies included | SD (TPR) | SD (FPR) | Corr. | Abu-Rumeileh | Leitão | Simon |
|---|---|---|---|---|---|---|---|
| Definite, probable, and possible sCJD cases | 1 | | | | × | | |
| **Male-to-female ratio >1** | | | | | | | |
| Definite sCJD cases | 0 | | | | | | |
| Definite and probable sCJD cases | 1 | | | | | | × |
| Definite, probable, and possible sCJD cases | 1 | | | | | | × |
| **Male-to-female ratio 0.75–1** | | | | | | | |
| Definite sCJD cases | 1 | | | | | × | |
| Definite and probable sCJD cases | 1 | | | | | × | |
| Definite, probable, and possible sCJD cases | 1 | | | | | × | |
| **Median age >65 years** | | | | | | | |
| Definite sCJD cases | 1 | | | | × | | |
| Definite and probable sCJD cases | 2 | 0 | 0.69 | NaN | × | | × |
| Definite, probable, and possible sCJD cases | 2 | 0 | 0.69 | NaN | × | | × |
| **Not only 14-3-3β investigated** | | | | | | | |
| Definite sCJD cases | 1 | | | | | × | |
| Definite and probable sCJD cases | 1 | | | | | × | |
| Definite, probable, and possible sCJD cases | 1 | | | | | × | |
| **RT-QuIC not investigated** | | | | | | | |
| Definite sCJD cases | 1 | | | | | × | |
| Definite and probable sCJD cases | 1 | | | | | × | |
| Definite, probable, and possible sCJD cases | 1 | | | | | × | |
| **Studies >2009** | | | | | | | |
| Definite sCJD cases | 2 | 0.58 | 0.99 | 1.00 | × | × | |
| Definite and probable sCJD cases | 3 | 0.42 | 0.85 | 0.61 | × | × | × |
| Definite, probable, and possible sCJD cases | 3 | 0.42 | 0.85 | 0.61 | × | × | × |
| **Studies with a priori cut-off** | | | | | | | |
| Definite sCJD cases | 1 | | | | | × | |
| Definite and probable sCJD cases | 2 | 0.63 | 0.23 | 1.00 | | × | × |
| Definite, probable, and possible sCJD cases | 2 | 0.63 | 0.23 | 1.00 | | × | × |
| **Studies with data-driven cut-off** | | | | | | | |

| Level of certainty of sCJD diagnosis | Studies included | SD (TPR) | SD (FPR) | Corr. | Abu-Rumeileh | Leitão | Simon |
|---|---|---|---|---|---|---|---|
| Definite sCJD cases | 1 | | | | × | | |
| Definite and probable sCJD cases | 1 | | | | × | | |
| Definite, probable, and possible sCJD cases | 1 | | | | × | | |
| **Study sample: only definite cases** | | | | | | | |
| Definite sCJD cases | 2 | 0.58 | 0.99 | 1.00 | × | × | |
| Definite and probable sCJD cases | 3 | 0.42 | 0.85 | 0.61 | × | × | × |
| Definite, probable, and possible sCJD cases | 3 | 0.42 | 0.85 | 0.61 | × | × | × |

**Table e-7: Results of meta-analyses involving RT-QuIC and subgroup analyses based on QUADAS-2 quality and clinical criteria**
Abbreviations of domains and criteria can be found in Table e-3.
FPR: false positive rate
NaN: not a number (correlation cannot be calculated)
SD: standard deviation
TPR: true positive rate
×: included in respective analysis

| Level of certainty of sCJD diagnosis | Studies included | SD (TPR) | SD (FPR) | Corr. | Abu-Rumeileh | Bizzi | Fiorini | Franceschini | Rhoads | Simon |
|---|---|---|---|---|---|---|---|---|---|---|
| **All studies** | | | | | | | | | | |
| Definite sCJD cases | 5 | 0.38 | 0.92 | 1.00 | × | × | × | × | × | |
| Definite and probable sCJD cases | 6 | 0.42 | 0.15 | 1.00 | × | × | × | × | × | × |
| Definite, probable, and possible sCJD cases | 6 | 0.42 | 0.15 | 1.00 | × | × | × | × | × | × |
| **Aβ42 not investigated** | | | | | | | | | | |
| Definite sCJD cases | 5 | 0.38 | 0.92 | 1.00 | × | × | × | × | × | |
| Definite and probable sCJD cases | 6 | 0.42 | 0.15 | 1.00 | × | × | × | × | × | × |
| Definite, probable, and possible sCJD cases | 6 | 0.42 | 0.15 | 1.00 | × | × | × | × | × | × |
| **Case-control studies** | | | | | | | | | | |
| Definite sCJD cases | 1 | | | | × | | | | | |
| Definite and probable sCJD cases | 1 | | | | × | | | | | |
| Definite, probable, and possible sCJD cases | 1 | | | | × | | | | | |
| **Cohort studies** | | | | | | | | | | |
| Definite sCJD cases | 4 | 0 | 0.00 | NaN | | × | × | × | × | |
| Definite and probable sCJD cases | 5 | 0.42 | 0.10 | 1.00 | | × | × | × | × | × |
| Definite, probable, and possible sCJD cases | 5 | 0.42 | 0.10 | 1.00 | | × | × | × | × | × |
| **D01 excl. high risk of bias (≥3 / 3 criteria)** | | | | | | | | | | |
| Definite sCJD cases | 4 | 0 | 0.00 | NaN | | × | × | × | × | |
| Definite and probable sCJD cases | 5 | 0.42 | 0.10 | 1.00 | | × | × | × | × | × |
| Definite, probable, and possible sCJD cases | 5 | 0.42 | 0.10 | 1.00 | | × | × | × | × | × |
| **D01 excl. high risk of bias (≥2 / 3 criteria)** | | | | | | | | | | |
| Definite sCJD cases | 4 | 0 | 0.00 | NaN | | × | × | × | × | |
| Definite and probable sCJD cases | 5 | 0.42 | 0.10 | 1.00 | | × | × | × | × | × |

| Level of certainty of sCJD diagnosis | Studies included | SD (TPR) | SD (FPR) | Corr. | Abu-Rumeileh | Bizzi | Fiorini | Franceschini | Rhoads | Simon |
|---|---|---|---|---|---|---|---|---|---|---|
| Definite, probable, and possible sCJD cases | 5 | 0.42 | 0.10 | 1.00 | | × | × | × | × | × |
| **D01 excl. high risk of bias (≥1 / 3 criteria)** | | | | | | | | | | |
| Definite sCJD cases | 3 | 0 | 0.00 | NaN | | | × | × | × | |
| Definite and probable sCJD cases | 4 | 0.19 | 0.11 | 1.00 | | | × | × | × | × |
| Definite, probable, and possible sCJD cases | 4 | 0.19 | 0.11 | 1.00 | | | × | × | × | × |
| **D02 excl. high risk of bias (≥1 / 2 criteria)** | | | | | | | | | | |
| Definite sCJD cases | 3 | 0 | 0.00 | NaN | | × | × | | × | |
| Definite and probable sCJD cases | 4 | 0.35 | 0.03 | 1.00 | | × | × | | × | × |
| Definite, probable, and possible sCJD cases | 4 | 0.35 | 0.03 | 1.00 | | × | × | | × | × |
| **D03 excl. high risk of bias (≥1 / 2 criteria)** | | | | | | | | | | |
| Definite sCJD cases | 5 | 0.38 | 0.92 | 1.00 | × | × | × | × | × | |
| Definite and probable sCJD cases | 5 | 0.46 | 1.17 | 1.00 | × | × | × | × | × | |
| Definite, probable, and possible sCJD cases | 5 | 0.46 | 1.17 | 1.00 | × | × | × | × | × | |
| **D04 excl. high risk of bias (≥3 / 4 criteria)** | | | | | | | | | | |
| Definite sCJD cases | 5 | 0.38 | 0.92 | 1.00 | × | × | × | × | × | |
| Definite and probable sCJD cases | 6 | 0.42 | 0.15 | 1.00 | × | × | × | × | × | × |
| Definite, probable, and possible sCJD cases | 6 | 0.42 | 0.15 | 1.00 | × | × | × | × | × | × |
| **D04 excl. high risk of bias (≥2 / 4 criteria)** | | | | | | | | | | |
| Definite sCJD cases | 5 | 0.38 | 0.92 | 1.00 | × | × | × | × | × | |
| Definite and probable sCJD cases | 6 | 0.42 | 0.15 | 1.00 | × | × | × | × | × | × |
| Definite, probable, and possible sCJD cases | 6 | 0.42 | 0.15 | 1.00 | × | × | × | × | × | × |
| **D04 excl. high risk of bias (≥1 / 4 criteria)** | | | | | | | | | | |
| Definite sCJD cases | 5 | 0.38 | 0.92 | 1.00 | × | × | × | × | × | |
| Definite and probable sCJD cases | 5 | 0.46 | 1.17 | 1.00 | × | × | × | × | × | |
| Definite, probable, and possible sCJD cases | 5 | 0.46 | 1.17 | 1.00 | × | × | × | × | × | |
| **D05 excl. poor applicability (≥1 / 3 criteria)** | | | | | | | | | | |
| Definite sCJD cases | 4 | 0.21 | 1.35 | 1.00 | × | | × | × | × | |
| Definite and probable sCJD cases | 5 | 0.22 | 0.16 | 1.00 | × | | × | × | × | × |
| Definite, probable, and possible sCJD cases | 5 | 0.22 | 0.16 | 1.00 | × | | × | × | × | × |
| **Disease duration not reported** | | | | | | | | | | |

| Level of certainty of sCJD diagnosis | Studies included | SD (TPR) | SD (FPR) | Corr. | Abu-Rumeileh | Bizzi | Fiorini | Franceschini | Rhoads | Simon |
|---|---|---|---|---|---|---|---|---|---|---|
| Definite sCJD cases | 1 | | | | | × | | | | |
| Definite and probable sCJD cases | 2 | 0.55 | 0.08 | -1.00 | | × | | | | × |
| Definite, probable, and possible sCJD cases | 2 | 0.55 | 0.08 | -1.00 | | × | | | | × |
| **Disease duration reported** | | | | | | | | | | |
| Definite sCJD cases | 4 | 0.21 | 1.35 | 1.00 | × | | × | × | × | |
| Definite and probable sCJD cases | 4 | 0.29 | 1.61 | 1.00 | × | | × | × | × | |
| Definite, probable, and possible sCJD cases | 4 | 0.29 | 1.61 | 1.00 | × | | × | × | × | |
| **K01 excl. high risk of bias** | | | | | | | | | | |
| Definite sCJD cases | 4 | 0 | 0.00 | NaN | | × | × | × | × | |
| Definite and probable sCJD cases | 5 | 0.42 | 0.10 | 1.00 | | × | × | × | × | × |
| Definite, probable, and possible sCJD cases | 5 | 0.42 | 0.10 | 1.00 | | × | × | × | × | × |
| **K01 excl. high/unclear risk of bias** | | | | | | | | | | |
| Definite sCJD cases | 4 | 0 | 0.00 | NaN | | × | × | × | × | |
| Definite and probable sCJD cases | 5 | 0.42 | 0.10 | 1.00 | | × | × | × | × | × |
| Definite, probable, and possible sCJD cases | 5 | 0.42 | 0.10 | 1.00 | | × | × | × | × | × |
| **K02 excl. high risk of bias** | | | | | | | | | | |
| Definite sCJD cases | 3 | 0 | 0.00 | NaN | | | × | × | × | |
| Definite and probable sCJD cases | 4 | 0.19 | 0.11 | 1.00 | | | × | × | × | × |
| Definite, probable, and possible sCJD cases | 4 | 0.19 | 0.11 | 1.00 | | | × | × | × | × |
| **K02 excl. high/unclear risk of bias** | | | | | | | | | | |
| Definite sCJD cases | 3 | 0 | 0.00 | NaN | | | × | × | × | |
| Definite and probable sCJD cases | 4 | 0.19 | 0.11 | 1.00 | | | × | × | × | × |
| Definite, probable, and possible sCJD cases | 4 | 0.19 | 0.11 | 1.00 | | | × | × | × | × |
| **K03 excl. high risk of bias** | | | | | | | | | | |
| Definite sCJD cases | 4 | 0 | 0.00 | NaN | | × | × | × | × | |
| Definite and probable sCJD cases | 5 | 0.42 | 0.10 | 1.00 | | × | × | × | × | × |
| Definite, probable, and possible sCJD cases | 5 | 0.42 | 0.10 | 1.00 | | × | × | × | × | × |
| **K03 excl. high/unclear risk of bias** | | | | | | | | | | |
| Definite sCJD cases | 3 | 0 | 0.00 | NaN | | × | | × | × | |
| Definite and probable sCJD cases | 4 | 0.43 | 0.06 | 1.00 | | × | | × | × | × |

| Level of certainty of sCJD diagnosis | Studies included | SD (TPR) | SD (FPR) | Corr. | Abu-Rumeileh | Bizzi | Fiorini | Franceschini | Rhoads | Simon |
|---|---|---|---|---|---|---|---|---|---|---|
| Definite, probable, and possible sCJD cases | 4 | 0.43 | 0.06 | 1.00 | | × | | × | × | × |
| **K04 excl. high risk of bias** | | | | | | | | | | |
| Definite sCJD cases | 4 | 0 | 0.00 | NaN | × | × | × | | × | |
| Definite and probable sCJD cases | 5 | 0.37 | 0.06 | 1.00 | × | × | × | | × | × |
| Definite, probable, and possible sCJD cases | 5 | 0.37 | 0.06 | 1.00 | × | × | × | | × | × |
| **K04 excl. high/unclear risk of bias** | | | | | | | | | | |
| Definite sCJD cases | 1 | | | | | | | | × | |
| Definite and probable sCJD cases | 1 | | | | | | | | × | |
| Definite, probable, and possible sCJD cases | 1 | | | | | | | | × | |
| **K05 excl. high risk of bias** | | | | | | | | | | |
| Definite sCJD cases | 4 | 0 | 0.00 | NaN | | × | × | × | × | |
| Definite and probable sCJD cases | 5 | 0.42 | 0.10 | 1.00 | | × | × | × | × | × |
| Definite, probable, and possible sCJD cases | 5 | 0.42 | 0.10 | 1.00 | | × | × | × | × | × |
| **K05 excl. high/unclear risk of bias** | | | | | | | | | | |
| Definite sCJD cases | 3 | 0.53 | 1.00 | 0.05 | | × | × | × | | |
| Definite and probable sCJD cases | 4 | 0.54 | 0.02 | -1.00 | | × | × | × | | × |
| Definite, probable, and possible sCJD cases | 4 | 0.54 | 0.02 | -1.00 | | × | × | × | | × |
| **K07 excl. high risk of bias** | | | | | | | | | | |
| Definite sCJD cases | 5 | 0.38 | 0.92 | 1.00 | × | × | × | × | × | |
| Definite and probable sCJD cases | 5 | 0.46 | 1.17 | 1.00 | × | × | × | × | × | |
| Definite, probable, and possible sCJD cases | 5 | 0.46 | 1.17 | 1.00 | × | × | × | × | × | |
| **K07 excl. high/unclear risk of bias** | | | | | | | | | | |
| Definite sCJD cases | 1 | | | | | | | × | | |
| Definite and probable sCJD cases | 1 | | | | | | | × | | |
| Definite, probable, and possible sCJD cases | 1 | | | | | | | × | | |
| **K08 excl. high risk of bias** | | | | | | | | | | |
| Definite sCJD cases | 5 | 0.38 | 0.92 | 1.00 | × | × | × | × | × | |
| Definite and probable sCJD cases | 6 | 0.42 | 0.15 | 1.00 | × | × | × | × | × | × |
| Definite, probable, and possible sCJD cases | 6 | 0.42 | 0.15 | 1.00 | × | × | × | × | × | × |
| **K09 excl. high risk of bias** | | | | | | | | | | |

| Level of certainty of sCJD diagnosis | Studies included | SD (TPR) | SD (FPR) | Corr. | Abu-Rumeileh | Bizzi | Fiorini | Franceschini | Rhoads | Simon |
|---|---|---|---|---|---|---|---|---|---|---|
| Definite sCJD cases | 5 | 0.38 | 0.92 | 1.00 | × | × | × | × | × | |
| Definite and probable sCJD cases | 5 | 0.46 | 1.17 | 1.00 | × | × | × | × | × | |
| Definite, probable, and possible sCJD cases | 5 | 0.46 | 1.17 | 1.00 | × | × | × | × | × | |
| **K09 excl. high/unclear risk of bias** | | | | | | | | | | |
| Definite sCJD cases | 5 | 0.38 | 0.92 | 1.00 | × | × | × | × | × | |
| Definite and probable sCJD cases | 5 | 0.46 | 1.17 | 1.00 | × | × | × | × | × | |
| Definite, probable, and possible sCJD cases | 5 | 0.46 | 1.17 | 1.00 | × | × | × | × | × | |
| **K10 excl. high risk of bias** | | | | | | | | | | |
| Definite sCJD cases | 5 | 0.38 | 0.92 | 1.00 | × | × | × | × | × | |
| Definite and probable sCJD cases | 6 | 0.42 | 0.15 | 1.00 | × | × | × | × | × | × |
| Definite, probable, and possible sCJD cases | 6 | 0.42 | 0.15 | 1.00 | × | × | × | × | × | × |
| **K10 excl. high/unclear risk of bias** | | | | | | | | | | |
| Definite sCJD cases | 5 | 0.38 | 0.92 | 1.00 | × | × | × | × | × | |
| Definite and probable sCJD cases | 6 | 0.42 | 0.15 | 1.00 | × | × | × | × | × | × |
| Definite, probable, and possible sCJD cases | 6 | 0.42 | 0.15 | 1.00 | × | × | × | × | × | × |
| **K11 excl. high/unclear risk of bias** | | | | | | | | | | |
| Definite sCJD cases | 5 | 0.38 | 0.92 | 1.00 | × | × | × | × | × | |
| Definite and probable sCJD cases | 6 | 0.42 | 0.15 | 1.00 | × | × | × | × | × | × |
| Definite, probable, and possible sCJD cases | 6 | 0.42 | 0.15 | 1.00 | × | × | × | × | × | × |
| **K12 excl. poor applicability** | | | | | | | | | | |
| Definite sCJD cases | 5 | 0.38 | 0.92 | 1.00 | × | × | × | × | × | |
| Definite and probable sCJD cases | 6 | 0.42 | 0.15 | 1.00 | × | × | × | × | × | × |
| Definite, probable, and possible sCJD cases | 6 | 0.42 | 0.15 | 1.00 | × | × | × | × | × | × |
| **K12 excl. poor/unclear applicability** | | | | | | | | | | |
| Definite sCJD cases | 5 | 0.38 | 0.92 | 1.00 | × | × | × | × | × | |
| Definite and probable sCJD cases | 5 | 0.46 | 1.17 | 1.00 | × | × | × | × | × | |
| Definite, probable, and possible sCJD cases | 5 | 0.46 | 1.17 | 1.00 | × | × | × | × | × | |
| **K13 excl. poor applicability** | | | | | | | | | | |
| Definite sCJD cases | 4 | 0.21 | 1.35 | 1.00 | × | | × | × | × | |
| Definite and probable sCJD cases | 5 | 0.22 | 0.16 | 1.00 | × | | × | × | × | × |

| Level of certainty of sCJD diagnosis | Studies included | SD (TPR) | SD (FPR) | Corr. | Abu-Rumeileh | Bizzi | Fiorini | Franceschini | Rhoads | Simon |
|---|---|---|---|---|---|---|---|---|---|---|
| Definite, probable, and possible sCJD cases | 5 | 0.22 | 0.16 | 1.00 | × | | × | × | × | × |
| **K13 excl. poor/unclear applicability** | | | | | | | | | | |
| Definite sCJD cases | 4 | 0.21 | 1.35 | 1.00 | × | | × | × | × | |
| Definite and probable sCJD cases | 5 | 0.22 | 0.16 | 1.00 | × | | × | × | × | × |
| Definite, probable, and possible sCJD cases | 5 | 0.22 | 0.16 | 1.00 | × | | × | × | × | × |
| **Male-to-female ratio <0.75** | | | | | | | | | | |
| Definite sCJD cases | 1 | | | | × | | | | | |
| Definite and probable sCJD cases | 1 | | | | × | | | | | |
| Definite, probable, and possible sCJD cases | 1 | | | | × | | | | | |
| **Male-to-female ratio >1** | | | | | | | | | | |
| Definite sCJD cases | 2 | 0 | 0.00 | NaN | | | × | | × | |
| Definite and probable sCJD cases | 3 | 0 | 0.00 | NaN | | | × | | × | × |
| Definite, probable, and possible sCJD cases | 3 | 0 | 0.00 | NaN | | | × | | × | × |
| **Male-to-female ratio 0.75–1** | | | | | | | | | | |
| Definite sCJD cases | 2 | 0.60 | 1.01 | -0.02 | | × | | × | | |
| Definite and probable sCJD cases | 2 | 0.68 | 0.82 | -0.29 | | × | | × | | |
| Definite, probable, and possible sCJD cases | 2 | 0.68 | 0.82 | -0.29 | | × | | × | | |
| **Median age >65 years** | | | | | | | | | | |
| Definite sCJD cases | 4 | 0.42 | 0.69 | 1.00 | × | × | | × | × | |
| Definite and probable sCJD cases | 5 | 0.44 | 0.09 | 1.00 | × | × | | × | × | × |
| Definite, probable, and possible sCJD cases | 5 | 0.44 | 0.09 | 1.00 | × | × | | × | × | × |
| **Studies >2009** | | | | | | | | | | |
| Definite sCJD cases | 5 | 0.38 | 0.92 | 1.00 | × | × | × | × | × | |
| Definite and probable sCJD cases | 6 | 0.42 | 0.15 | 1.00 | × | × | × | × | × | × |
| Definite, probable, and possible sCJD cases | 6 | 0.42 | 0.15 | 1.00 | × | × | × | × | × | × |
| **Studies with a priori cut-off** | | | | | | | | | | |
| Definite sCJD cases | 4 | 0 | 0.00 | NaN | | × | × | × | × | |
| Definite and probable sCJD cases | 5 | 0.42 | 0.10 | 1.00 | | × | × | × | × | × |
| Definite, probable, and possible sCJD cases | 5 | 0.42 | 0.10 | 1.00 | | × | × | × | × | × |
| **Study sample: not only definite cases** | | | | | | | | | | |

| Level of certainty of sCJD diagnosis | Studies included | SD (TPR) | SD (FPR) | Corr. | Abu-Rumeileh | Bizzi | Fiorini | Franceschini | Rhoads | Simon |
|---|---|---|---|---|---|---|---|---|---|---|
| Definite sCJD cases | 2 | 0 | 0.02 | NaN | | | × | × | | |
| Definite and probable sCJD cases | 2 | 0 | 0.01 | NaN | | | × | × | | |
| Definite, probable, and possible sCJD cases | 2 | 0 | 0.01 | NaN | | | × | × | | |
| **Study sample: only definite cases** | | | | | | | | | | |
| Definite sCJD cases | 3 | 0.26 | 0.11 | 1.00 | × | × | | | × | |
| Definite and probable sCJD cases | 4 | 0.36 | 0.02 | 1.00 | × | × | | | × | × |
| Definite, probable, and possible sCJD cases | 4 | 0.36 | 0.02 | 1.00 | × | × | | | × | × |

**Table e-8: Results of meta-analyses involving S100B and subgroup analyses based on QUADAS-2 quality and clinical criteria**
Abbreviations of domains and criteria can be found in Table e-3.
FPR: false positive rate
NaN: not a number (correlation cannot be calculated)
SD: standard deviation
TPR: true positive rate
×: included in respective analysis

| Level of certainty of sCJD diagnosis | Studies included | SD (TPR) | SD (FPR) | Corr. | Baldeiras | Chohan | Sanchez-Juan |
|---|---|---|---|---|---|---|---|
| **All studies** | | | | | | | |
| Definite sCJD cases | 2 | 0.91 | 0.17 | 1.00 | × | × | |
| Definite and probable sCJD cases | 2 | 0.91 | 0.17 | 1.00 | × | × | |
| Definite, probable, and possible sCJD cases | 3 | 0.64 | 0.01 | 1.00 | × | × | × |
| **Aβ42 investigated** | | | | | | | |
| Definite sCJD cases | 1 | | | | × | | |
| Definite and probable sCJD cases | 1 | | | | × | | |
| Definite, probable, and possible sCJD cases | 1 | | | | × | | |
| **Aβ42 not investigated** | | | | | | | |
| Definite sCJD cases | 1 | | | | | × | |
| Definite and probable sCJD cases | 1 | | | | | × | |
| Definite, probable, and possible sCJD cases | 2 | 0.43 | 0.60 | -1.00 | | × | × |
| **Case-control studies** | | | | | | | |
| Definite sCJD cases | 1 | | | | × | | |
| Definite and probable sCJD cases | 1 | | | | × | | |
| Definite, probable, and possible sCJD cases | 1 | | | | × | | |
| **Cohort studies** | | | | | | | |
| Definite sCJD cases | 1 | | | | | × | |
| Definite and probable sCJD cases | 1 | | | | | × | |
| Definite, probable, and possible sCJD cases | 2 | 0.43 | 0.60 | -1.00 | | × | × |
| **D01 excl. high risk of bias (≥3 / 3 criteria)** | | | | | | | |
| Definite sCJD cases | 1 | | | | | × | |
| Definite and probable sCJD cases | 1 | | | | | × | |

| Level of certainty of sCJD diagnosis | Studies included | SD (TPR) | SD (FPR) | Corr. | Baldeiras | Chohan | Sanchez-Juan |
|---|---|---|---|---|---|---|---|
| Definite, probable, and possible sCJD cases | 2 | 0.43 | 0.60 | -1.00 | | × | × |
| **D01 excl. high risk of bias (≥2 / 3 criteria)** | | | | | | | |
| Definite sCJD cases | 1 | | | | | × | |
| Definite and probable sCJD cases | 1 | | | | | × | |
| Definite, probable, and possible sCJD cases | 2 | 0.43 | 0.60 | -1.00 | | × | × |
| **D02 excl. high risk of bias (≥1 / 2 criteria)** | | | | | | | |
| Definite sCJD cases | 1 | | | | | × | |
| Definite and probable sCJD cases | 1 | | | | | × | |
| Definite, probable, and possible sCJD cases | 2 | 0.43 | 0.60 | -1.00 | | × | × |
| **D03 excl. high risk of bias (≥1 / 2 criteria)** | | | | | | | |
| Definite sCJD cases | 1 | | | | × | | |
| Definite and probable sCJD cases | 1 | | | | × | | |
| Definite, probable, and possible sCJD cases | 2 | 0.18 | 0.70 | 1.00 | × | | × |
| **D04 excl. high risk of bias (≥3 / 4 criteria)** | | | | | | | |
| Definite sCJD cases | 2 | 0.91 | 0.17 | 1.00 | × | × | |
| Definite and probable sCJD cases | 2 | 0.91 | 0.17 | 1.00 | × | × | |
| Definite, probable, and possible sCJD cases | 3 | 0.64 | 0.01 | 1.00 | × | × | × |
| **D04 excl. high risk of bias (≥2 / 4 criteria)** | | | | | | | |
| Definite sCJD cases | 1 | | | | | × | |
| Definite and probable sCJD cases | 1 | | | | | × | |
| Definite, probable, and possible sCJD cases | 2 | 0.43 | 0.60 | -1.00 | | × | × |
| **D05 excl. poor applicability (≥1 / 3 criteria)** | | | | | | | |
| Definite sCJD cases | 1 | | | | × | | |
| Definite and probable sCJD cases | 1 | | | | × | | |
| Definite, probable, and possible sCJD cases | 2 | 0.18 | 0.70 | 1.00 | × | | × |
| **Disease duration not reported** | | | | | | | |
| Definite sCJD cases | 1 | | | | | × | |
| Definite and probable sCJD cases | 1 | | | | | × | |
| Definite, probable, and possible sCJD cases | 2 | 0.43 | 0.60 | -1.00 | | × | × |
| **Disease duration reported** | | | | | | | |

| Level of certainty of sCJD diagnosis | Studies included | SD (TPR) | SD (FPR) | Corr. | Baldeiras | Chohan | Sanchez-Juan |
|---|---|---|---|---|---|---|---|
| Definite sCJD cases | 1 | | | | × | | |
| Definite and probable sCJD cases | 1 | | | | × | | |
| Definite, probable, and possible sCJD cases | 1 | | | | × | | |
| **K01 excl. high risk of bias** | | | | | | | |
| Definite sCJD cases | 1 | | | | | × | |
| Definite and probable sCJD cases | 1 | | | | | × | |
| Definite, probable, and possible sCJD cases | 1 | | | | | × | |
| **K01 excl. high/unclear risk of bias** | | | | | | | |
| Definite sCJD cases | 1 | | | | | × | |
| Definite and probable sCJD cases | 1 | | | | | × | |
| Definite, probable, and possible sCJD cases | 1 | | | | | × | |
| **K02 excl. high risk of bias** | | | | | | | |
| Definite sCJD cases | 1 | | | | | × | |
| Definite and probable sCJD cases | 1 | | | | | × | |
| Definite, probable, and possible sCJD cases | 2 | 0.43 | 0.60 | -1.00 | | × | × |
| **K03 excl. high risk of bias** | | | | | | | |
| Definite sCJD cases | 0 | | | | | | |
| Definite and probable sCJD cases | 0 | | | | | | |
| Definite, probable, and possible sCJD cases | 1 | | | | | | × |
| **K04 excl. high risk of bias** | | | | | | | |
| Definite sCJD cases | 2 | 0.91 | 0.17 | 1.00 | × | × | |
| Definite and probable sCJD cases | 2 | 0.91 | 0.17 | 1.00 | × | × | |
| Definite, probable, and possible sCJD cases | 3 | 0.64 | 0.01 | 1.00 | × | × | × |
| **K04 excl. high/unclear risk of bias** | | | | | | | |
| Definite sCJD cases | 1 | | | | | × | |
| Definite and probable sCJD cases | 1 | | | | | × | |
| Definite, probable, and possible sCJD cases | 1 | | | | | × | |
| **K05 excl. high risk of bias** | | | | | | | |
| Definite sCJD cases | 1 | | | | | × | |
| Definite and probable sCJD cases | 1 | | | | | × | |

| Level of certainty of sCJD diagnosis | Studies included | SD (TPR) | SD (FPR) | Corr. | Baldeiras | Chohan | Sanchez-Juan |
|---|---|---|---|---|---|---|---|
| Definite, probable, and possible sCJD cases | 2 | 0.43 | 0.60 | -1.00 | | × | × |
| **K05 excl. high/unclear risk of bias** | | | | | | | |
| Definite sCJD cases | 1 | | | | | × | |
| Definite and probable sCJD cases | 1 | | | | | × | |
| Definite, probable, and possible sCJD cases | 2 | 0.43 | 0.60 | -1.00 | | × | × |
| **K07 excl. high risk of bias** | | | | | | | |
| Definite sCJD cases | 1 | | | | × | | |
| Definite and probable sCJD cases | 1 | | | | × | | |
| Definite, probable, and possible sCJD cases | 2 | 0.18 | 0.70 | 1.00 | × | | × |
| **K08 excl. high risk of bias** | | | | | | | |
| Definite sCJD cases | 2 | 0.91 | 0.17 | 1.00 | × | × | |
| Definite and probable sCJD cases | 2 | 0.91 | 0.17 | 1.00 | × | × | |
| Definite, probable, and possible sCJD cases | 3 | 0.64 | 0.01 | 1.00 | × | × | × |
| **K09 excl. high risk of bias** | | | | | | | |
| Definite sCJD cases | 1 | | | | | × | |
| Definite and probable sCJD cases | 1 | | | | | × | |
| Definite, probable, and possible sCJD cases | 2 | 0.43 | 0.60 | -1.00 | | × | × |
| **K09 excl. high/unclear risk of bias** | | | | | | | |
| Definite sCJD cases | 1 | | | | | × | |
| Definite and probable sCJD cases | 1 | | | | | × | |
| Definite, probable, and possible sCJD cases | 2 | 0.43 | 0.60 | -1.00 | | × | × |
| **K11 excl. high/unclear risk of bias** | | | | | | | |
| Definite sCJD cases | 1 | | | | | × | |
| Definite and probable sCJD cases | 1 | | | | | × | |
| Definite, probable, and possible sCJD cases | 2 | 0.43 | 0.60 | -1.00 | | × | × |
| **K12 excl. poor applicability** | | | | | | | |
| Definite sCJD cases | 1 | | | | × | | |
| Definite and probable sCJD cases | 1 | | | | × | | |
| Definite, probable, and possible sCJD cases | 2 | 0.18 | 0.70 | 1.00 | × | | × |
| **K12 excl. poor/unclear applicability** | | | | | | | |

| Level of certainty of sCJD diagnosis | Studies included | SD (TPR) | SD (FPR) | Corr. | Baldeiras | Chohan | Sanchez-Juan |
|---|---|---|---|---|---|---|---|
| Definite sCJD cases | 0 | | | | | | |
| Definite and probable sCJD cases | 0 | | | | | | |
| Definite, probable, and possible sCJD cases | 1 | | | | | | × |
| **K13 excl. poor applicability** | | | | | | | |
| Definite sCJD cases | 2 | 0.91 | 0.17 | 1.00 | × | × | |
| Definite and probable sCJD cases | 2 | 0.91 | 0.17 | 1.00 | × | × | |
| Definite, probable, and possible sCJD cases | 3 | 0.64 | 0.01 | 1.00 | × | × | × |
| **K13 excl. poor/unclear applicability** | | | | | | | |
| Definite sCJD cases | 2 | 0.91 | 0.17 | 1.00 | × | × | |
| Definite and probable sCJD cases | 2 | 0.91 | 0.17 | 1.00 | × | × | |
| Definite, probable, and possible sCJD cases | 3 | 0.64 | 0.01 | 1.00 | × | × | × |
| **Male-to-female ratio >1** | | | | | | | |
| Definite sCJD cases | 1 | | | | | × | |
| Definite and probable sCJD cases | 1 | | | | | × | |
| Definite, probable, and possible sCJD cases | 1 | | | | | × | |
| **Male-to-female ratio 0.75–1** | | | | | | | |
| Definite sCJD cases | 1 | | | | × | | |
| Definite and probable sCJD cases | 1 | | | | × | | |
| Definite, probable, and possible sCJD cases | 1 | | | | × | | |
| **Median age >65 years** | | | | | | | |
| Definite sCJD cases | 2 | 0.91 | 0.17 | 1.00 | × | × | |
| Definite and probable sCJD cases | 2 | 0.91 | 0.17 | 1.00 | × | × | |
| Definite, probable, and possible sCJD cases | 2 | 0.91 | 0.17 | 1.00 | × | × | |
| **Studies <2009** | | | | | | | |
| Definite sCJD cases | 2 | 0.91 | 0.17 | 1.00 | × | × | |
| Definite and probable sCJD cases | 2 | 0.91 | 0.17 | 1.00 | × | × | |
| Definite, probable, and possible sCJD cases | 3 | 0.64 | 0.01 | 1.00 | × | × | × |
| **Studies with a priori cut-off** | | | | | | | |
| Definite sCJD cases | 1 | | | | | × | |
| Definite and probable sCJD cases | 1 | | | | | × | |

| Level of certainty of sCJD diagnosis | Studies included | SD (TPR) | SD (FPR) | Corr. | Baldeiras | Chohan | Sanchez-Juan |
|---|---|---|---|---|---|---|---|
| Definite, probable, and possible sCJD cases | 2 | 0.43 | 0.60 | -1.00 | | × | × |
| **Studies with data-driven cut-off** | | | | | | | |
| Definite sCJD cases | 1 | | | | × | | |
| Definite and probable sCJD cases | 1 | | | | × | | |
| Definite, probable, and possible sCJD cases | 1 | | | | × | | |
| **Study sample: not only definite cases** | | | | | | | |
| Definite sCJD cases | 1 | | | | | × | |
| Definite and probable sCJD cases | 1 | | | | | × | |
| Definite, probable, and possible sCJD cases | 2 | 0.43 | 0.60 | -1.00 | | × | × |
| **Study sample: only definite cases** | | | | | | | |
| Definite sCJD cases | 1 | | | | × | | |
| Definite and probable sCJD cases | 1 | | | | × | | |
| Definite, probable, and possible sCJD cases | 1 | | | | × | | |

**Table e-9: Results of meta-analyses involving t-tau and subgroup analyses based on QUADAS-2 quality and clinical criteria**
Abbreviations of domains and criteria can be found in Table e-3.
FPR: false positive rate
NaN: not a number (correlation cannot be calculated)
SD: standard deviation
TPR: true positive rate
×: included in respective analysis

| **Level of certainty of sCJD diagnosis** | **Studies included** | **SD (TPR)** | **SD (FPR)** | **Corr.** | **Abu-Rumeileh** | **Baldeiras** | **Bizzi** | **Bongianni** | **Chohan** | **Fiorini** | **Franceschini** | **Lattanzio** | **Otto** | **Sanchez-Juan** | **Simon** | **Van Everbroeck** |
|---|---|---|---|---|---|---|---|---|---|---|---|---|---|---|---|---|
| **All studies** | | | | | | | | | | | | | | | | |
| Definite, probable, and possible sCJD cases | 12 | 0.32 | 1.00 | 0.10 | × | × | × | × | × | × | × | × | × | × | × | × |
| Definite and probable sCJD cases | 11 | 0.35 | 1.01 | 0.22 | × | × | × | × | × | × | × | × | × | | × | × |
| Definite sCJD cases | 9 | 0.38 | 0.74 | 0.41 | × | × | × | × | × | × | × | × | × | | | |
| **Aβ42 investigated** | | | | | | | | | | | | | | | | |
| Definite, probable, and possible sCJD cases | 2 | 0 | 0.83 | NaN | | × | | | | | | × | | | | |
| Definite and probable sCJD cases | 2 | 0.01 | 0.83 | 1.00 | | × | | | | | | × | | | | |
| Definite sCJD cases | 2 | 0 | 0.83 | NaN | | × | | | | | | × | | | | |
| **Aβ42 not investigated** | | | | | | | | | | | | | | | | |
| Definite, probable, and possible sCJD cases | 10 | 0.36 | 1.02 | 0.09 | × | | × | × | × | × | × | | × | × | × | × |
| Definite and probable sCJD cases | 9 | 0.40 | 1.04 | 0.21 | × | | × | × | × | × | × | | × | | × | × |
| Definite sCJD cases | 7 | 0.46 | 0.67 | 0.38 | × | | × | × | × | × | × | | × | | | |
| **Case-control studies** | | | | | | | | | | | | | | | | |
| Definite, probable, and possible sCJD cases | 3 | 0.44 | 1.35 | 1.00 | × | × | | × | | | | | | | | |
| Definite and probable sCJD cases | 3 | 0.44 | 1.35 | 1.00 | × | × | | × | | | | | | | | |
| Definite sCJD cases | 3 | 0.54 | 1.35 | 1.00 | × | × | | × | | | | | | | | |
| **Cohort studies** | | | | | | | | | | | | | | | | |
| Definite, probable, and possible sCJD cases | 9 | 0.31 | 0.91 | -0.18 | | | × | | × | × | × | × | × | × | × | × |
| Definite and probable sCJD cases | 8 | 0.36 | 0.92 | -0.08 | | | × | | × | × | × | × | × | | × | × |
| Definite sCJD cases | 6 | 0.34 | 0.51 | -0.09 | | | × | | × | × | × | × | × | | | |
| **D01 excl. high risk of bias (≥3 / 3 criteria)** | | | | | | | | | | | | | | | | |
| Definite, probable, and possible sCJD cases | 9 | 0.31 | 0.91 | -0.18 | | | × | | × | × | × | × | × | × | × | × |
| Definite and probable sCJD cases | 8 | 0.36 | 0.92 | -0.08 | | | × | | × | × | × | × | × | | × | × |

| Level of certainty of sCJD diagnosis | Studies included | SD (TPR) | SD (FPR) | Corr. | Abu-Rumeileh | Baldeiras | Bizzi | Bongianni | Chohan | Fiorini | Franceschini | Lattanzio | Otto | Sanchez-Juan | Simon | Van Everbroeck |
|---|---|---|---|---|---|---|---|---|---|---|---|---|---|---|---|---|
| Definite sCJD cases | 6 | 0.34 | 0.51 | -0.09 | | | × | | × | × | × | × | × | | | |
| **D01 excl. high risk of bias (≥2 / 3 criteria)** | | | | | | | | | | | | | | | | |
| Definite, probable, and possible sCJD cases | 9 | 0.31 | 0.91 | -0.18 | | | × | | × | × | × | × | × | × | × | × |
| Definite and probable sCJD cases | 8 | 0.36 | 0.92 | -0.08 | | | × | | × | × | × | × | × | | × | × |
| Definite sCJD cases | 6 | 0.34 | 0.51 | -0.09 | | | × | | × | × | × | × | × | | | |
| **D01 excl. high risk of bias (≥1 / 3 criteria)** | | | | | | | | | | | | | | | | |
| Definite, probable, and possible sCJD cases | 4 | 0.13 | 0.52 | 0.27 | | | | | | × | × | × | | | × | |
| Definite and probable sCJD cases | 4 | 0.09 | 0.53 | 0.30 | | | | | | × | × | × | | | × | |
| Definite sCJD cases | 3 | 0.09 | 0.09 | -1.00 | | | | | | × | × | × | | | | |
| **D02 excl. high risk of bias (≥1 / 2 criteria)** | | | | | | | | | | | | | | | | |
| Definite, probable, and possible sCJD cases | 7 | 0.02 | 1.21 | 1.00 | | | × | × | × | × | | | | × | × | × |
| Definite and probable sCJD cases | 6 | 0.21 | 1.25 | 0.39 | | | × | × | × | × | | | | | × | × |
| Definite sCJD cases | 4 | 0.17 | 0.59 | -1.00 | | | × | × | × | × | | | | | | |
| **D03 excl. high risk of bias (≥1 / 2 criteria)** | | | | | | | | | | | | | | | | |
| Definite, probable, and possible sCJD cases | 10 | 0.24 | 1.12 | 0.16 | × | × | × | × | | × | × | × | × | × | | × |
| Definite and probable sCJD cases | 9 | 0.24 | 1.14 | 0.71 | × | × | × | × | | × | × | × | × | | | × |
| Definite sCJD cases | 8 | 0.36 | 0.79 | 1.00 | × | × | × | × | | × | × | × | × | | | |
| **D04 excl. high risk of bias (≥3 / 4 criteria)** | | | | | | | | | | | | | | | | |
| Definite, probable, and possible sCJD cases | 12 | 0.32 | 1.00 | 0.10 | × | × | × | × | × | × | × | × | × | × | × | × |
| Definite and probable sCJD cases | 11 | 0.35 | 1.01 | 0.22 | × | × | × | × | × | × | × | × | × | | × | × |
| Definite sCJD cases | 9 | 0.38 | 0.74 | 0.41 | × | × | × | × | × | × | × | × | × | | | |
| **D04 excl. high risk of bias (≥2 / 4 criteria)** | | | | | | | | | | | | | | | | |
| Definite, probable, and possible sCJD cases | 11 | 0.32 | 0.96 | 0.08 | × | | × | × | × | × | × | × | × | × | × | × |
| Definite and probable sCJD cases | 10 | 0.36 | 0.97 | 0.20 | × | | × | × | × | × | × | × | × | | × | × |
| Definite sCJD cases | 8 | 0.39 | 0.58 | 0.35 | × | | × | × | × | × | × | × | × | | | |
| **D04 excl. high risk of bias (≥1 / 4 criteria)** | | | | | | | | | | | | | | | | |
| Definite, probable, and possible sCJD cases | 5 | 0.32 | 0.39 | 0.94 | × | | × | × | | × | × | | | | | |
| Definite and probable sCJD cases | 5 | 0.32 | 0.39 | 0.94 | × | | × | × | | × | × | | | | | |
| Definite sCJD cases | 5 | 0 | 0.34 | NaN | × | | × | × | | × | × | | | | | |
| **D05 excl. poor applicability (≥1 / 3 criteria)** | | | | | | | | | | | | | | | | |

| **Level of certainty of sCJD diagnosis** | **Studies included** | **SD (TPR)** | **SD (FPR)** | **Corr.** | **Abu-Rumeileh** | **Baldeiras** | **Bizzi** | **Bongianni** | **Chohan** | **Fiorini** | **Franceschini** | **Lattanzio** | **Otto** | **Sanchez-Juan** | **Simon** | **Van Everbroeck** |
|---|---|---|---|---|---|---|---|---|---|---|---|---|---|---|---|---|
| Definite, probable, and possible sCJD cases | 10 | 0.28 | 1.06 | 0.08 | × | × | | × | | × | × | × | × | × | × | × |
| Definite and probable sCJD cases | 9 | 0.25 | 1.10 | 0.52 | × | × | | × | | × | × | × | × | | × | × |
| Definite sCJD cases | 7 | 0.39 | 0.82 | 1.00 | × | × | | × | | × | × | × | × | | | |
| **Disease duration not reported** | | | | | | | | | | | | | | | | |
| Definite, probable, and possible sCJD cases | 5 | 0.35 | 0.58 | 0.28 | | | × | | × | | | | × | × | × | |
| Definite and probable sCJD cases | 4 | 0.44 | 0.60 | 0.43 | | | × | | × | | | | × | | × | |
| Definite sCJD cases | 3 | 0.46 | 0.65 | 0.29 | | | × | | × | | | | × | | | |
| **Disease duration reported** | | | | | | | | | | | | | | | | |
| Definite, probable, and possible sCJD cases | 7 | 0.21 | 1.25 | 0.18 | × | × | | × | | × | × | × | | | | × |
| Definite and probable sCJD cases | 7 | 0.23 | 1.25 | 0.16 | × | × | | × | | × | × | × | | | | × |
| Definite sCJD cases | 6 | 0.29 | 0.80 | 1.00 | × | × | | × | | × | × | × | | | | |
| **K01 excl. high risk of bias** | | | | | | | | | | | | | | | | |
| Definite, probable, and possible sCJD cases | 8 | 0.35 | 0.92 | -0.07 | | | × | | × | × | × | × | × | | × | × |
| Definite and probable sCJD cases | 8 | 0.36 | 0.92 | -0.08 | | | × | | × | × | × | × | × | | × | × |
| Definite sCJD cases | 6 | 0.34 | 0.51 | -0.09 | | | × | | × | × | × | × | × | | | |
| **K01 excl. high/unclear risk of bias** | | | | | | | | | | | | | | | | |
| Definite, probable, and possible sCJD cases | 6 | 0.31 | 0.56 | -0.28 | | | × | | × | × | × | × | | | × | |
| Definite and probable sCJD cases | 6 | 0.32 | 0.56 | -0.27 | | | × | | × | × | × | × | | | × | |
| Definite sCJD cases | 5 | 0.21 | 0.41 | -1.00 | | | × | | × | × | × | × | | | | |
| **K02 excl. high risk of bias** | | | | | | | | | | | | | | | | |
| Definite, probable, and possible sCJD cases | 7 | 0.30 | 0.94 | -0.53 | | | | | × | × | × | × | | × | × | × |
| Definite and probable sCJD cases | 6 | 0.37 | 0.95 | -0.47 | | | | | × | × | × | × | | | × | × |
| Definite sCJD cases | 4 | 0.36 | 0.39 | -1.00 | | | | | × | × | × | × | | | | |
| **K02 excl. high/unclear risk of bias** | | | | | | | | | | | | | | | | |
| Definite, probable, and possible sCJD cases | 4 | 0.13 | 0.52 | 0.27 | | | | | | × | × | × | | | × | |
| Definite and probable sCJD cases | 4 | 0.09 | 0.53 | 0.30 | | | | | | × | × | × | | | × | |
| Definite sCJD cases | 3 | 0.09 | 0.09 | -1.00 | | | | | | × | × | × | | | | |
| **K03 excl. high risk of bias** | | | | | | | | | | | | | | | | |
| Definite, probable, and possible sCJD cases | 7 | 0.27 | 0.59 | 0.35 | | | × | | | × | × | × | × | × | × | |
| Definite and probable sCJD cases | 6 | 0.24 | 0.59 | 1.00 | | | × | | | × | × | × | × | | × | |

| **Level of certainty of sCJD diagnosis** | **Studies included** | **SD (TPR)** | **SD (FPR)** | **Corr.** | **Abu-Rumeileh** | **Baldeiras** | **Bizzi** | **Bongianni** | **Chohan** | **Fiorini** | **Franceschini** | **Lattanzio** | **Otto** | **Sanchez-Juan** | **Simon** | **Van Everbroeck** |
|---|---|---|---|---|---|---|---|---|---|---|---|---|---|---|---|---|
| Definite sCJD cases | 5 | 0.24 | 0.48 | 1.00 | | | × | | | × | × | × | × | | | |
| **K03 excl. high/unclear risk of bias** | | | | | | | | | | | | | | | | |
| Definite, probable, and possible sCJD cases | 5 | 0.24 | 0.64 | 1.00 | | | × | | | | × | × | × | | × | |
| Definite and probable sCJD cases | 5 | 0.25 | 0.64 | 1.00 | | | × | | | | × | × | × | | × | |
| Definite sCJD cases | 4 | 0.26 | 0.55 | 1.00 | | | × | | | | × | × | × | | | |
| **K04 excl. high risk of bias** | | | | | | | | | | | | | | | | |
| Definite, probable, and possible sCJD cases | 9 | 0.15 | 1.13 | 0.29 | × | × | × | × | × | × | | | | × | × | × |
| Definite and probable sCJD cases | 8 | 0.25 | 1.16 | 0.43 | × | × | × | × | × | × | | | | | × | × |
| Definite sCJD cases | 6 | 0.31 | 0.88 | 0.26 | × | × | × | × | × | × | | | | | | |
| **K04 excl. high/unclear risk of bias** | | | | | | | | | | | | | | | | |
| Definite, probable, and possible sCJD cases | 2 | 0 | 1.11 | NaN | | | | × | × | | | | | | | |
| Definite and probable sCJD cases | 2 | 0 | 1.11 | NaN | | | | × | × | | | | | | | |
| Definite sCJD cases | 2 | 0.12 | 1.12 | 1.00 | | | | × | × | | | | | | | |
| **K05 excl. high risk of bias** | | | | | | | | | | | | | | | | |
| Definite, probable, and possible sCJD cases | 10 | 0.32 | 1.03 | 0.09 | | | × | × | × | × | × | × | × | × | × | × |
| Definite and probable sCJD cases | 9 | 0.37 | 1.04 | 0.21 | | | × | × | × | × | × | × | × | | × | × |
| Definite sCJD cases | 7 | 0.38 | 0.64 | 0.32 | | | × | × | × | × | × | × | × | | | |
| **K05 excl. high/unclear risk of bias** | | | | | | | | | | | | | | | | |
| Definite, probable, and possible sCJD cases | 10 | 0.32 | 1.03 | 0.09 | | | × | × | × | × | × | × | × | × | × | × |
| Definite and probable sCJD cases | 9 | 0.37 | 1.04 | 0.21 | | | × | × | × | × | × | × | × | | × | × |
| Definite sCJD cases | 7 | 0.38 | 0.64 | 0.32 | | | × | × | × | × | × | × | × | | | |
| **K07 excl. high risk of bias** | | | | | | | | | | | | | | | | |
| Definite, probable, and possible sCJD cases | 10 | 0.24 | 1.12 | 0.16 | × | × | × | × | | × | × | × | × | × | | × |
| Definite and probable sCJD cases | 9 | 0.24 | 1.14 | 0.71 | × | × | × | × | | × | × | × | × | | | × |
| Definite sCJD cases | 8 | 0.36 | 0.79 | 1.00 | × | × | × | × | | × | × | × | × | | | |
| **K07 excl. high/unclear risk of bias** | | | | | | | | | | | | | | | | |
| Definite, probable, and possible sCJD cases | 4 | 0.45 | 0.85 | 0.89 | | | | × | | | × | × | × | | | |
| Definite and probable sCJD cases | 4 | 0.44 | 0.86 | 0.94 | | | | × | | | × | × | × | | | |
| Definite sCJD cases | 4 | 0.49 | 0.84 | 1.00 | | | | × | | | × | × | × | | | |
| **K08 excl. high risk of bias** | | | | | | | | | | | | | | | | |

| Level of certainty of sCJD diagnosis | Studies included | SD (TPR) | SD (FPR) | Corr. | Abu-Rumeileh | Baldeiras | Bizzi | Bongianni | Chohan | Fiorini | Franceschini | Lattanzio | Otto | Sanchez-Juan | Simon | Van Everbroeck |
|---|---|---|---|---|---|---|---|---|---|---|---|---|---|---|---|---|
| Definite, probable, and possible sCJD cases | 12 | 0.32 | 1.00 | 0.10 | × | × | × | × | × | × | × | × | × | × | × | × |
| Definite and probable sCJD cases | 11 | 0.35 | 1.01 | 0.22 | × | × | × | × | × | × | × | × | × | | × | × |
| Definite sCJD cases | 9 | 0.38 | 0.74 | 0.41 | × | × | × | × | × | × | × | × | × | | | |
| **K08 excl. high/unclear risk of bias** | | | | | | | | | | | | | | | | |
| Definite, probable, and possible sCJD cases | 1 | | | | | | | × | | | | | | | | |
| Definite and probable sCJD cases | 1 | | | | | | | × | | | | | | | | |
| Definite sCJD cases | 1 | | | | | | | × | | | | | | | | |
| **K09 excl. high risk of bias** | | | | | | | | | | | | | | | | |
| Definite, probable, and possible sCJD cases | 9 | 0.36 | 1.08 | 0.02 | × | | × | × | × | × | × | | × | × | | × |
| Definite and probable sCJD cases | 8 | 0.41 | 1.09 | 0.13 | × | | × | × | × | × | × | | × | | | × |
| Definite sCJD cases | 7 | 0.46 | 0.67 | 0.38 | × | | × | × | × | × | × | | × | | | |
| **K09 excl. high/unclear risk of bias** | | | | | | | | | | | | | | | | |
| Definite, probable, and possible sCJD cases | 9 | 0.36 | 1.08 | 0.02 | × | | × | × | × | × | × | | × | × | | × |
| Definite and probable sCJD cases | 8 | 0.41 | 1.09 | 0.13 | × | | × | × | × | × | × | | × | | | × |
| Definite sCJD cases | 7 | 0.46 | 0.67 | 0.38 | × | | × | × | × | × | × | | × | | | |
| **K10 excl. high risk of bias** | | | | | | | | | | | | | | | | |
| Definite, probable, and possible sCJD cases | 7 | 0.24 | 0.63 | 1.00 | × | | × | × | | × | × | × | | | × | |
| Definite and probable sCJD cases | 7 | 0.25 | 0.63 | 1.00 | × | | × | × | | × | × | × | | | × | |
| Definite sCJD cases | 6 | 0 | 0.29 | NaN | × | | × | × | | × | × | × | | | | |
| **K10 excl. high/unclear risk of bias** | | | | | | | | | | | | | | | | |
| Definite, probable, and possible sCJD cases | 7 | 0.24 | 0.63 | 1.00 | × | | × | × | | × | × | × | | | × | |
| Definite and probable sCJD cases | 7 | 0.25 | 0.63 | 1.00 | × | | × | × | | × | × | × | | | × | |
| Definite sCJD cases | 6 | 0 | 0.29 | NaN | × | | × | × | | × | × | × | | | | |
| **K11 excl. high/unclear risk of bias** | | | | | | | | | | | | | | | | |
| Definite, probable, and possible sCJD cases | 11 | 0.32 | 0.96 | 0.08 | × | | × | × | × | × | × | × | × | × | × | × |
| Definite and probable sCJD cases | 10 | 0.36 | 0.97 | 0.20 | × | | × | × | × | × | × | × | × | | × | × |
| Definite sCJD cases | 8 | 0.39 | 0.58 | 0.35 | × | | × | × | × | × | × | × | × | | | |
| **K12 excl. poor applicability** | | | | | | | | | | | | | | | | |
| Definite, probable, and possible sCJD cases | 11 | 0.25 | 1.06 | 0.26 | × | × | × | × | | × | × | × | × | × | × | × |
| Definite and probable sCJD cases | 10 | 0.22 | 1.09 | 0.98 | × | × | × | × | | × | × | × | × | | × | × |

| Level of certainty of sCJD diagnosis | Studies included | SD (TPR) | SD (FPR) | Corr. | Abu-Rumeileh | Baldeiras | Bizzi | Bongianni | Chohan | Fiorini | Franceschini | Lattanzio | Otto | Sanchez-Juan | Simon | Van Everbroeck |
|---|---|---|---|---|---|---|---|---|---|---|---|---|---|---|---|---|
| Definite sCJD cases | 8 | 0.36 | 0.79 | 1.00 | × | × | × | × | | × | × | × | × | | | |
| **K12 excl. poor/unclear applicability** | | | | | | | | | | | | | | | | |
| Definite, probable, and possible sCJD cases | 8 | 0.23 | 0.97 | -0.27 | × | | × | | | × | × | × | × | × | | × |
| Definite and probable sCJD cases | 7 | 0.22 | 0.96 | 0.21 | × | | × | | | × | × | × | × | | | × |
| Definite sCJD cases | 6 | 0.24 | 0.43 | 1.00 | × | | × | | | × | × | × | × | | | |
| **K13 excl. poor applicability** | | | | | | | | | | | | | | | | |
| Definite, probable, and possible sCJD cases | 11 | 0.35 | 0.99 | 0.03 | × | × | | × | × | × | × | × | × | × | × | × |
| Definite and probable sCJD cases | 10 | 0.39 | 1.01 | 0.14 | × | × | | × | × | × | × | × | × | | × | × |
| Definite sCJD cases | 8 | 0.43 | 0.74 | 0.36 | × | × | | × | × | × | × | × | × | | | |
| **K13 excl. poor/unclear applicability** | | | | | | | | | | | | | | | | |
| Definite, probable, and possible sCJD cases | 11 | 0.35 | 0.99 | 0.03 | × | × | | × | × | × | × | × | × | × | × | × |
| Definite and probable sCJD cases | 10 | 0.39 | 1.01 | 0.14 | × | × | | × | × | × | × | × | × | | × | × |
| Definite sCJD cases | 8 | 0.43 | 0.74 | 0.36 | × | × | | × | × | × | × | × | × | | | |
| **Male-to-female ratio <0.75** | | | | | | | | | | | | | | | | |
| Definite, probable, and possible sCJD cases | 2 | 0.10 | 0.21 | 1.00 | × | | | | | | | | × | | | |
| Definite and probable sCJD cases | 2 | 0.10 | 0.21 | 1.00 | × | | | | | | | | × | | | |
| Definite sCJD cases | 2 | 0.10 | 0.21 | 1.00 | × | | | | | | | | × | | | |
| **Male-to-female ratio >1** | | | | | | | | | | | | | | | | |
| Definite, probable, and possible sCJD cases | 3 | 0.32 | 0.43 | 0.50 | | | | | × | × | | | | | × | |
| Definite and probable sCJD cases | 3 | 0.32 | 0.43 | 0.50 | | | | | × | × | | | | | × | |
| Definite sCJD cases | 2 | 0.40 | 0.41 | -1.00 | | | | | × | × | | | | | | |
| **Male-to-female ratio 0.75–1** | | | | | | | | | | | | | | | | |
| Definite, probable, and possible sCJD cases | 5 | 0.27 | 0.97 | 1.00 | | × | × | × | | | × | × | | | | |
| Definite and probable sCJD cases | 5 | 0.31 | 0.97 | 1.00 | | × | × | × | | | × | × | | | | |
| Definite sCJD cases | 5 | 0.24 | 0.99 | 1.00 | | × | × | × | | | × | × | | | | |
| **Median age >65 years** | | | | | | | | | | | | | | | | |
| Definite, probable, and possible sCJD cases | 9 | 0.41 | 0.79 | 0.36 | × | × | × | × | × | | × | × | × | | × | |
| Definite and probable sCJD cases | 9 | 0.41 | 0.79 | 0.36 | × | × | × | × | × | | × | × | × | | × | |
| Definite sCJD cases | 8 | 0.40 | 0.82 | 0.48 | × | × | × | × | × | | × | × | × | | | |
| **RT-QuIC not investigated** | | | | | | | | | | | | | | | | |

| Level of certainty of sCJD diagnosis | Studies included | SD (TPR) | SD (FPR) | Corr. | Abu-Rumeileh | Baldeiras | Bizzi | Bongianni | Chohan | Fiorini | Franceschini | Lattanzio | Otto | Sanchez-Juan | Simon | Van Everbroeck |
|---|---|---|---|---|---|---|---|---|---|---|---|---|---|---|---|---|
| Definite, probable, and possible sCJD cases | 5 | 0.33 | 0.65 | 0.20 | | × | | | × | | | | × | × | | × |
| Definite and probable sCJD cases | 4 | 0.45 | 0.69 | 0.18 | | × | | | × | | | | × | | | × |
| Definite sCJD cases | 3 | 0.55 | 0.19 | 1.00 | | × | | | × | | | | × | | | |
| **Studies <2009** | | | | | | | | | | | | | | | | |
| Definite, probable, and possible sCJD cases | 5 | 0.33 | 0.65 | 0.20 | | × | | | × | | | | × | × | | × |
| Definite and probable sCJD cases | 4 | 0.45 | 0.69 | 0.18 | | × | | | × | | | | × | | | × |
| Definite sCJD cases | 3 | 0.55 | 0.19 | 1.00 | | × | | | × | | | | × | | | |
| **Studies >2009** | | | | | | | | | | | | | | | | |
| Definite, probable, and possible sCJD cases | 7 | 0.24 | 0.63 | 1.00 | × | | × | × | | × | × | × | | | × | |
| Definite and probable sCJD cases | 7 | 0.25 | 0.63 | 1.00 | × | | × | × | | × | × | × | | | × | |
| Definite sCJD cases | 6 | 0 | 0.29 | NaN | × | | × | × | | × | × | × | | | | |
| **Studies with a priori cut-off** | | | | | | | | | | | | | | | | |
| Definite, probable, and possible sCJD cases | 8 | 0.26 | 1.16 | -0.08 | | | × | × | × | × | × | | | × | × | × |
| Definite and probable sCJD cases | 7 | 0.33 | 1.18 | 0.03 | | | × | × | × | × | × | | | | × | × |
| Definite sCJD cases | 5 | 0.18 | 0.52 | -1.00 | | | × | × | × | × | × | | | | | |
| **Studies with data-driven cut-off** | | | | | | | | | | | | | | | | |
| Definite, probable, and possible sCJD cases | 4 | 0.24 | 0.48 | 1.00 | × | × | | | | | | × | × | | | |
| Definite and probable sCJD cases | 4 | 0.20 | 0.48 | 1.00 | × | × | | | | | | × | × | | | |
| Definite sCJD cases | 4 | 0.27 | 0.47 | 1.00 | × | × | | | | | | × | × | | | |
| **Study sample: not only definite cases** | | | | | | | | | | | | | | | | |
| Definite, probable, and possible sCJD cases | 6 | 0.38 | 0.49 | -0.43 | | | | | × | × | × | × | × | × | | |
| Definite and probable sCJD cases | 5 | 0.46 | 0.46 | -0.32 | | | | | × | × | × | × | × | | | |
| Definite sCJD cases | 5 | 0.41 | 0.46 | -0.36 | | | | | × | × | × | × | × | | | |
| **Study sample: only definite cases** | | | | | | | | | | | | | | | | |
| Definite, probable, and possible sCJD cases | 6 | 0.21 | 1.38 | 1.00 | × | × | × | × | | | | | | | × | × |
| Definite and probable sCJD cases | 6 | 0.21 | 1.38 | 1.00 | × | × | × | × | | | | | | | × | × |
| Definite sCJD cases | 4 | 0.42 | 1.17 | 1.00 | × | × | × | × | | | | | | | | |
